# Being responsible for COPD - lung disease as a manifestation of structural violence

**DOI:** 10.1101/2023.09.06.23295021

**Authors:** Parris J Williams, Sara C Buttery, Anthony A Laverty, Nicholas S Hopkinson

## Abstract

Lung health, the development of lung disease, and how well a person with lung disease is able to live, all depend on a wide range of societal factors. Considering COPD as a manifestation of structural violence, something that continues to be done to people, despite it being largely preventable, makes the causal processes more apparent and the responsibility to interrupt or alleviate these clearer. We developed a taxonomy to describe this, containing five domains. 1)Avoidable lung harms: (i)processes impacting on lung development (ii)processes which disadvantage lung health in particular groups across the life course. 2)Diagnostic Delay: (i)healthcare factors (ii)norms and attitudes that mean that COPD is not diagnosed in a timely way, denying people with COPD effective treatment. 3)Inadequate COPD Care: ways in which the provision of care for people with COPD falls short of what is needed to ensure that they are able to enjoy the best possible health, considered as (i)healthcare resource allocation (ii)norms and attitudes influencing clinical practice. 4)Low status of COPD: ways in which both COPD as a condition and people with COPD are held in less regard and considered less of a priority than other comparable health problems. 5)Lack of Support: factors that make living with COPD more difficult than it should be (i)socioenvironmental factors (ii)factors that promote social isolation. This model has relevance for policymakers, healthcare professionals and the public as an educational resource, to change clinical practices and priorities and to stimulate advocacy and activism with the goal of the elimination of COPD.

## INTRODUCTION

Chronic obstructive pulmonary disease (COPD) is one of the leading causes of disability and premature death. The link between socioeconomic deprivation and health status is undeniable, and in common with all long-term health conditions, COPD is not distributed equally within and between populations. Healthcare systems and healthcare professionals tend to view health conditions as something that people “have”. In this paper, using the concept of structural violence, we argue that lung disease, as exemplified by COPD, can also usefully be represented as something that has been “done to” people. By framing it in this way, the causal processes become more apparent and with that, the need for means to interrupt or alleviate these mechanisms becomes clearer.

The term structural violence was coined in 1969 by Johan Galtung, who defined violence as present “when human beings are being influenced so that their actual somatic and mental realizations are below their potential realizations”(1). Violence can be direct and physical, or structural. This is analogous to Stokely Carmichael’s distinction between two types of racism: individual racism and institutional racism(2). The absence of direct violence is peace, the absence of structural violence is social justice(1). A key consideration is that structural violence considers influences that limit individuals’ *potential* realisations. That is to say, life experiences that are actually possible to achieve. For example, aging is inevitable, but the circumstances and manner in which we age vary enormously impacting on individual wellbeing and indeed the possibility of living a life of adequate length.

The use of the term “structural violence”, which is both more direct than usual discourse while also unfamiliar but easily understood, is intended to focus attention on the way in which structural factors in societies both cause people to develop COPD who need not have done so, and disadvantage people who have COPD so that their lives are more limited than they could be. Focus on these is needed to develop strategies and solutions to reduce or eliminate the burden of COPD.

We therefore aimed to develop a taxonomy of structural violence, using an iterative process to refine the structural social factors that bear on COPD, which people working in this field can build on to guide research, advocacy and reform.

## MODEL DEVELOPMENT

The model development followed a partnership-based approach, with collaboration between the research team, experts in various aspects of lung health and people affected by COPD. It was informed by the Six Steps in Quality Development framework (6SQuID), in particular (i) defining and understanding the problem and its causes; (ii) identifying which causal or contextual factors are modifiable: which have the greatest scope for change and who would benefit most; (iii) deciding on mechanisms of change(3).

Following informal discussions with respiratory experts and considering responses to Asthma + Lung UK (A+LUK) surveys of people with respiratory disease(4, 5), the authors proposed an initial model containing five domains of concern: avoidable risks that cause COPD, diagnostic delay, inadequate COPD care, low status of COPD, lack of support that people living with COPD experience(6). These categories were then circulated online to an expert panel, using SurveyMonkey (Momentive Inc, San Mateo, California, USA) to canvas opinion about the validity of this structure, as well as to rate the importance of key elements that should be contained in each domain and suggest additional ones. Expert input through this process led to an increased emphasis on early life and transgenerational factors as well as the impact of racism.

The survey was also sent to members of an A+LUK patient group who ranked elements to confirm their importance and were able to offer additional possible items. This led to an increased emphasis on the impact of stigma from the public and healthcare system as well as demands for legislation to tackle avoidable lung harms such as poverty and air pollution. Further stakeholder meetings were held to review and refine the model and evidence base allowing a deeper discussion around key concepts. Further details of the development are available in an online supplement.

Where mechanisms are described in the model they are in general only cited once, in what was deemed to be the most relevant domain, based on an assessment of impact or causal priority. For example, poor housing is a risk factor for developing lung disease but also makes living with lung disease more difficult.

### A CONCEPTUAL MODEL OF COPD AS A MANIFESTATION OF STRUCTURAL VIOLENCE

The final model included the following five domains, each consisting of two sub-domains (Figure 1).

**Figure 1:**
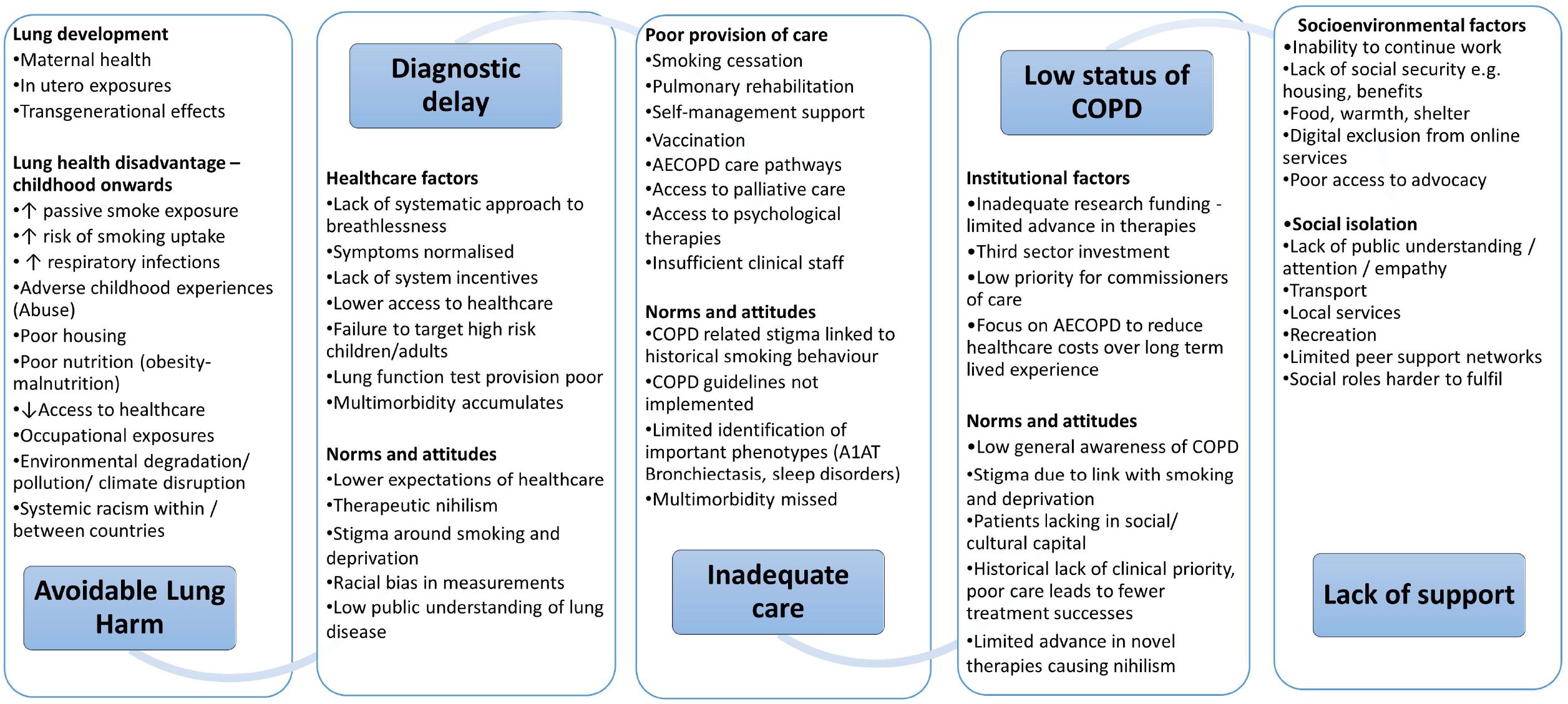
COPD as a locus for structural violence.

Avoidable lung harms: (i) Processes that impact on lung development (ii) Processes which disadvantage lung health in particular groups across the life course.

Diagnostic Delay: comprising (i) Healthcare factors and (ii) Norms and attitudes that mean that COPD is not diagnosed in a timely way, denying people with COPD effective treatment to improve/maintain quality of life and improve prognosis.

Inadequate COPD Care: ways in which the provision of care for people with COPD falls short of what is needed to ensure that they are able to enjoy the best possible health, considered as (i) Healthcare resource allocation (ii) Norms and attitudes influencing clinical practice.

Low status of COPD: ways in which both COPD as a condition and people with COPD are held in less regard and considered less of a priority than for other health problems (i) Institutional factors (ii) Norms and attitudes.

Lack of Support: factors that make living with COPD more difficult than it should be. These are categorised as (i) Socioenvironmental factors (ii) Factors that promote social isolation.

### Avoidable Lung Harms

The lung pathology that underpins COPD - airway inflammation and remodelling, bronchiolitis, alveolar destruction, and pulmonary vascular damage - occurs firstly, because of factors that interfere with lung growth both before and after birth and across generations. An adverse fetal environment produces lung structural changes, altered immunology, low birthweight and prematurity, sensitization to later insults and premature aging (telomere shortening)(7) as well as increasing cardiovascular risk driving multimorbidity(8). The second main factor is exposures that accelerate lung function decline across the life course(9, 10).

COPD is closely tied to inequality and deprivation(7, 11, 12), and in particular failure to ensure the health and wellbeing of mothers and children(13). As Michael Marmot has observed: “Child poverty is a political choice”. Absolute and relative poverty reduction(14, 15), tackling corporate determinants of health like the relative expense of healthy diets vs poor ones(16) and the promotion of alcohol(17), addressing neighbourhood and partner violence(18), structured early years support such as the US Head Start(19) or the UK’s SureStart schemes(20), policies that reduce workplace stress and occupational exposures(21), as well as reducing indoor and outdoor air pollution(22) and implementing comprehensive tobacco control measures(23) would all reduce the avoidable harm done to children’s lungs and thus reduce their risk of developing COPD in later life.

Most of the structural factors that drive lung health disadvantage in early life continue to operate across the life course. Failure to address smoking at the time of delivery increases children’s exposure to environmental tobacco smoke, and smoking behaviour is passed on to children by carers and peer groups(24), sustaining health inequality. Smoking is the leading preventable cause of premature death and disability, responsible for half the difference in life expectancy between rich and poor(25), while two thirds of people who continue to smoke will die from a smoking-related disease(26). Childhood smoking uptake substantially increases the risk of lung disease(27). There has been a failure to take action proportionate to this knowledge. In the UK, for example, there was a 36-year gap between the 1962 publication of *Smoking Kills* by the Royal College of Physicians and the first comprehensive government tobacco control plan in 1998. Smoking rates in UK 15-year-olds fell from 20% to 3% in the following 20 years, having been unchanged at that level for 20 years prior. There is no technical reason why the measures to reduce the affordability, availability and appeal of smoking could not have been taken earlier, and responsibility for millions of children taking up smoking rests with adult policy-makers. Globally, although there has been progress since the WHO’s Framework Convention on Tobacco Control treaty was instituted, virtually no country has chosen to implement the full range of measures needed to achieve its objectives(23, 28).

As with smoking, efforts to prevent youth uptake of vaping by reducing its affordability, availability, and appeal(29) to non-smokers continue to be inadequate to the task(28, 30). Illicit drug use, both inhaled and intravenous is another cause of COPD(31) that is determined by modifiable social factors and amenable to poverty reduction and other social interventions.

Respiratory infections in early childhood are associated with increased risk of death in adult life, from respiratory causes including COPD(32), while tuberculosis is also an important and socially determined cause of COPD(9). Modifiable factors that increase the risk of childhood respiratory infections and their severity include poor housing and nutrition, reduced access to healthcare, smoking, and exposure to indoor and outdoor air pollution.

Occupations where workers are exposed to dust fumes and chemicals increase the risk of developing obstructive lung disease(9, 21). Structural issues here include the lower socioeconomic status of factory and cleaning work and decisions about whether or not to provide workers with appropriate protection(33).

Global heating, climate disruption and air pollution cause COPD through a range of synergistic mechanisms including increased lung toxicity from particulates and NOx, as well as increased allergen exposure driving asthma incidence(34, 35). Active transport and clean public transportation reduce air pollution, while physical inactivity is associated with accelerated lung function decline, so failure to facilitate and promote physical activity is also a systemic risk(36).

### Diagnostic delay

The diagnostic criteria for COPD are straightforward, but the condition is typically diagnosed when there is already substantial lung damage and often when there have been suggestive symptoms for several years, meaning that patients miss out on interventions that could improve quality of life, delay progression and improve survival(37-40). Healthcare factors include a lack of systematic approach to breathlessness in midlife, with symptoms normalised as an expected consequence of aging or a “smoker’s cough”(38, 41). Allowing slowly progressive breathlessness to restrict daily physical activity levels may also promote multimorbidity, as sedentarism increase the risk of other long-term conditions including diabetes, hypertension and osteoporosis(36, 42). There is typically a lack of system incentives to identify COPD, lung function testing provision is poor and bias in measurement may systematically under-diagnose lung disease in some ethnic groups. High-risk groups such as those with a history of childhood chest disease or those in occupations associated with inhaled exposures or high smoking rates, those with mental health problems and the homeless are likely to be groups that also have less access to healthcare and lower expectations of healthcare.

There is generally poor awareness of COPD and lung disease and there has been a lack of strategies to communicate this and change health behaviour(43). People with symptoms may consider them self-inflicted, worry about stigma or worry that symptoms represent cancer. Therapeutic nihilism and stigma around COPD produce a self-fulfilling failure to make the diagnosis(44, 45).

### Inadequate care

COPD management aims to improve breathlessness and other symptoms, reduce disease progression and improve prognosis(46, 47). The UK’s National Institute for Health and Care Excellence (NICE) has defined “Five Fundamentals of COPD Care”: smoking cessation, vaccination, pulmonary rehabilitation, personalised self-management plans, and optimising treatment for multi-morbidity(46). However, most people with COPD are currently denied effective treatment both in low- and high-income countries(4, 48-50). Limited time and resources mean that basics like inhaler technique are not delivered. An insufficient respiratory workforce(39) and the move away from specialist respiratory care can also mean that treatable phenotypes including bronchiectasis, sleep-disordered breathing and pulmonary hypertension may go unidentified and untreated. The acceptance of symptoms as normal also means that multi-morbidity including cardiac disease, lung cancer and mental health problems go underdiagnosed and undertreated(6). Referral for specialist intervention like lung volume reduction procedures often occurs too late, if considered at all(51). Despite high symptom burden, COPD patient access to palliative care is limited(52, 53), as is access to treatment for psychological comorbidities(54). Care pathways do not systematise universal engagement with charities that could support patients(39).

Globally, the majority of people with COPD, are drawn from among the lowest income people in low-income countries, and do not have access to affordable medication(55) in addition, access to smoking cessation pharmacotherapy is also limited(56) and research and development focus on high income countries.

The explanations for poor care can be found in an interplay between healthcare system choices about prioritisation and norms and attitudes of healthcare workers and patients(57, 58), which drive a mutually reinforcing therapeutic nihilism(47).

### Low status of COPD

In 2019, there were estimated to be 212 million people diagnosed with COPD globally, and COPD accounted for 3.3 million deaths(10). Given its impacts on both healthcare systems and the lives of individual citizens, COPD receives proportionately less attention and investment in clinical care and research(39, 59) than would be expected(11). This is matched by a lack of public awareness of COPD as a health condition(60, 61). Respiratory disease is often omitted from healthcare policy discussion, even when highly relevant(62). Choices about which conditions to link to performance targets have impacted on COPD care. In the UK for example, National Service Frameworks, 10-year plans to achieve performance targets for conditions including cancer, coronary heart disease and stroke, accidents and mental health, were introduced around the turn of the century. The omission of respiratory disease from these has inevitably led to a diversion of resources and attention towards the conditions prioritised.

In addition, meagre allocation of research funds has translated into a lack of progress in understanding COPD and in the development of new treatments, while this lack of progress has deterred further investment(63). The low status of COPD clinical care also makes COPD research more difficult, as the starting point of a well-phenotyped patient established on standard care is less accessible than for example a person with coronary artery disease, where treatment is typically highly protocolised and relatively well-resourced.

The socioeconomic distribution of COPD means that people with the condition lack social and cultural capital which impacts on how they are perceived by healthcare professionals. Stigma is an important aspect of living with COPD, feeding into the low priority that the condition and people with it are given(44, 45, 58).

Care pathways for AECOPD have changed little over recent decades, in stark contrast to the treatment of myocardial infarction and stroke. Although mortality and readmission rates after hospitalisation for AECOPD are high, acceptance of this as a normal aspect of COPD means that post-discharge support and investigation is typically limited(64).

### Lack of support for living with COPD

The experience of having COPD is influenced for the worse by factors that make living with the condition more difficult. COPD is more common in poorer people and increases the likelihood that people will have to give up work prematurely, reducing their income and savings(65). Lack of money, poor housing and absent transport deny people with COPD the chance to mitigate some of the limitations caused by the disease, and living in a cold damp home increases the risk of acute exacerbations(66). Choices about welfare such as disability benefit and public provision of resources such as public transport, libraries, clubs will influence the constraints which poorer individuals with COPD live under(14). Older people with COPD may lack digital access or literacy, so digital by default services become inaccessible(67).

Stigma and a lack of public understanding make living with COPD more difficult. Related to but distinct from the more quantitative domain of social welfare, are factors that determine the experience of social isolation and loneliness including limited peer support networks, inability to fulfil social roles such as grandparenting and participation in social activities and recreation(68).

### Mitigating structural violence - scope and scale

Having set out the mechanisms through which structural violence manifests in COPD, the question arises as to what should be done about them? For healthcare professionals these can be considered in terms of what can be addressed by making systematic improvements to clinical practice, by advocacy and by healthcare professionals collectively through representative bodies such as Thoracic Societies (Table 1).

**Table 1.** Responsibility to address structural violence related to COPD.

|  |  |
| --- | --- |
| Policy makers | • Prioritise child and maternal wellbeing<br>• Adopt health in all policies<br>• Justice as fairness - policies that achieve redistribution and reparation<br>• Address intersectional injustices around ethnicity, gender, disability<br>• Legislate to protect citizens<br>• Address commercial determinants of health<br>• Enforce protections for people<br>• Provide equivalent resources to address respiratory conditions |
| The Public | • Acknowledge long term health conditions as structurally determined<br>• Become more aware of respiratory disease<br>• Demand policy makers address structural issues |
| People with COPD | • Self-advocacy<br>• Demand policy makers address structural issues |
| Healthcare professionals as individuals | • Clinical care<br>• Address stigma<br>• Clinical guidelines<br>• Promote awareness of lung disease in colleagues and public<br>• Advocacy<br>• Ensure fair commissioning of care for respiratory conditions<br>• Demand policy makers address structural issues |

### COPD and legal considerations

A person with lung disease may have been the victim of one of three broad types of injustice with possible legal remedies (Figure 2). Firstly, an individual or entity may have harmed them directly by exposing them to noxious materials. The tobacco industry is the obvious example, but other polluters or occupational exposures are also relevant. The second broad possibility is that an individual or entity with a duty to protect them from harm has failed to do this. In the case of government this could represent a failure to legislate, to comply with legislation or to enforce legislation. Examples include inaction on air quality and failure to restrain the harm caused by the tobacco industry(23). The third potential injustice is the failure to provide effective treatments or the means to access them. Inadequate provision of the Five Fundamentals of COPD care, including pulmonary rehabilitation and smoking cessation services is an example of this(46).

**Figure 2:**
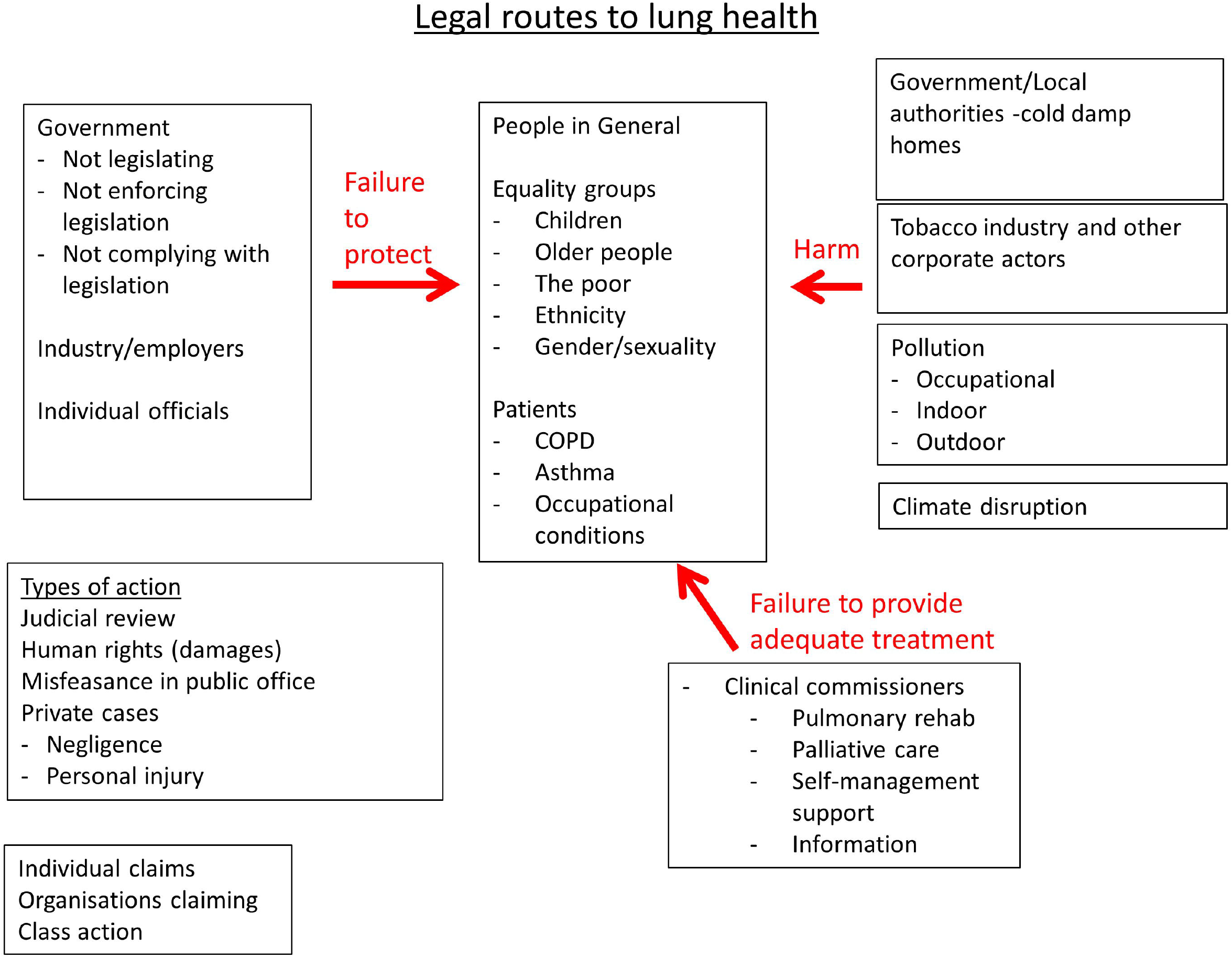
Legal routes to lung health.

The existence of these injustices raises the possibility that there might be legal remedies. The practical implications of this will vary from country to country depending on legal systems, but potential approaches depend on the entity considered to be at fault. There may also be specific issues where the group affected come from a designated group or minority. For government or quasi-governmental bodies judicial review and human rights-based approaches might be considered. Private actions for negligence or personal injury may be appropriate in some situations. Possible claims might be from individuals or class actions. Exemplary cases may prompt action. In 2020 Ella Kissi-Debra, a 9-year-old girl who died in 2013 became the first person in the world to have air pollution given as a cause of death. Her case has been a spur to expand London’s Ultra Low Emission Zone which is intended to limit the most polluting vehicles from driving in the city(69).

A more comprehensive table setting out evidence for harms related to each domain and potential mitigation is provided in the Online Supplement.

## DISCUSSION

The model described in this paper was developed through an iterative process involving a range of experts and people with COPD. Its five domains provide a systematic approach to considering the factors that cause people to develop COPD and to live lives that are limited in terms of wellbeing and agency – the extent to which they are able to pursue goals and objectives that they have reason to value. The model should help guide advocacy, modify clinical priorities and practice, and be a stimulus to activism for people with COPD and those who care about them.

Within each domain we set out the key processes that are in play, presenting salient examples and potential strategies for mitigation (see online Table E1). The model is intended to serve as a starting point; eliminating structural violence requires action at a range of scales.

In three of the five domains, we identified a subset of structural factors that could be defined as representing “norms and attitudes”. Although these are to a great extent culturally and institutionally determined, they nevertheless are likely to be the most amenable to individual, smaller-scale and local action by healthcare professionals.

Potential action ranges from steps taken to ensure that health literacy is addressed, and encouragement given to engage with health charities in all individual patient consultations, through decisions to provide an appropriate proportion of healthcare resources to the needs of people with COPD, through societal choices to reduce or eliminate poverty and inequality.

Much of the literature in this field comes from high income countries, which itself is a manifestation of structural injustice(70). Most of the items discussed are likely to apply across different countries and we use examples from a range of regions. The persistence of structural factors that cause and worsen COPD in wealthy countries with well-developed healthcare systems is striking, though the very existence of low-income countries can be considered to represent injustice and structural violence on a much greater scale.

A key underpinning element of structural violence is the impact of anthropogenic global heating. This will increase the occurrence of lung disease, make living with lung disease more difficult and reduce the resilience of societies in general and healthcare systems specifically(71). The fact that the countries and people most vulnerable to the effects of climate disruption are precisely those least responsible for it should be a readily accessible analogy for the processes addressed in this paper - people with COPD and their families are typically from social groups least likely to set laws or determine the behaviour of corporations.

Like any model, the 5 domains of structural violence and the elements they contain, may be subject to revision. We believe that the categories identified by our consensus process will prove useful and they are also intended to be as unobtrusive as possible, so that the content rather than the classification is the focus.

A criticism of this model is that it displaces concepts of personal responsibility and agency. Do people who make bad choices deserve to experience bad outcomes? There are a number of arguments against this. First, the approaches are not mutually exclusive. People can be taken to be free to choose within the situation they find themselves, but the point of structural violence as a concept is precisely that the range of choices available to an individual is limited by the structural processes that we describe. Second, individual choice is of no relevance to processes acting in early life and before birth – welfare policies that have the intent of chastening the undeserving poor, have the consequence of blighting the lives of the innocent unborn. In particular, choice is of limited relevance to smoking, which is driven for most people by an addiction starting in childhood and as such is better thought of as a decades-long failure of adults to protect children.

There are other economic arguments that addressing structural violence is a general good – investment in public health is likely to improve economic performance, a general benefit that goes beyond just those who are prevented from developing COPD or where the impact of the condition is reduced(72). Many of the interventions that would reduce the incidence of COPD would also reduce other long-term conditions and improve the general population’s experience of fairness and other aspects of quality of life.

### Conclusion

As Virchow famously asserted “Medicine is a social science and politics is nothing else but medicine on a large scale. Medicine as a social science, as the science of human beings, has the obligation to point out problems and to attempt their theoretical solution”(73). The model developed and presented in this paper, highlights how chronic obstructive pulmonary disease is a manifestation of structural violence and argues that rather than observing COPD as something people have done to themselves, we should view it as something that has been and continues to be done to them. This model has relevance for policymakers, healthcare professionals and the public as an education resource, to change clinical practices and priorities and to stimulate advocacy and activism for social justice and reparations.

## CONTRIBUTIONS

The initial model was developed by PJW, SCB. AAL and NSH. The collection and collation of stakeholder input was conducted by PJW. NSH wrote the first draft, to which all authors contributed and approved the final version.

## Data Availability

We do not have consent to share survey data collected by Asthma + Lung UK.

## ACKNOWLEDGEMENTS

We are very grateful for the support, input and insight from people with COPD who participated in the project, as well as to the following clinical and academic colleagues for their suggestions and help in the development process.

Mona Bafadhel, King’s College London

Andrew Bush, Imperial College London

James Dodd, University of Bristol

Rachael Evans, University of Leicester

John Hurst, University College, London

Rasleen Kahai, Royal Brompton and Harefield Hospitals

Anu Kumar, NHS City & Hackney Clinical Commissioning Group

Keir Lewis, Hywel Dda University Health Board

John Moxham, King’s College London

Najib Rahman, Oxford University

Victoria Singh, Royal Brompton and Harefield Hospitals

Ian Sinha, University of Liverpool

Laura-Jane Smith, King’s College London

Paul Walker, British Thoracic Society / University of Liverpool

Andrew Whittamore, Asthma + Lung UK

Sarah Woolnough, Asthma + Lung UK

## Notes

Conflicts of interest; NSH is Chair of Action on Smoking and Health and Medical Director of Asthma + Lung UK. AAL is a Trustee of Action on Smoking and Health. Other authors have no conflict of interest to declare.

### Competing Interest Statement

The authors have declared no competing interest.

### Funding Statement

This work did not receive any specific funding.

### Author Declarations

Participants completing the questionnaire consented to their anonymised data being used for research purposes and ethical approval for this analysis was granted by Imperial College Research and Integrity Team (IREC; 20IC6625).

