## Supplementary material for "Being responsible for COPD - lung disease as a manifestation of structural violence": Online Supplement

^1^Parris J Williams, ^1^Sara C Buttery, ^2^Anthony A Laverty, ^1^Nicholas S Hopkinson

^1^National Heart and Lung Institute, Imperial College

^2^Public Health Policy Evaluation Unit, School of Public Health, Imperial College London, London, UK

Professor Nicholas S. Hopkinson (Corresponding author)

National Heart and Lung Institute, Imperial College,

Royal Brompton Hospital Campus,

Fulham Road, London SW3 6HP

**Contents:**

| 2 | Model development – extended methods |
| --- | --- |
| 4 | Survey results COPD experts |
| 4 | Table E2: Expert panel demographics |
| 5 | Figures E1-5 Rating of factors contributing to different domains (COPD experts) |
| 10 | Survey results: Asthma + Lung UK patient panel |
| 10 | Table E3. Asthma+Lung UK supporter base demographics |
| 11 | Figures E6-10 Rating of factors contributing to different domains (COPD patients) |
| 16 | Mitigation strategies |
| 18 | Table E4. Mitigation categories and quotations |
| 19 | Structural violence Final model |
| 20 | Table E5: Structural violence: examples mechanisms and mitigations |

**MODEL DEVELOPMENT**

The model development followed a partnership-based approach, with collaboration between the research team, experts in various aspects of lung health and people affected by COPD. It was informed by the Six Steps in Quality Development framework (6SQuID), in particular (i) defining and understanding the problem and its causes; (ii) identifying which causal or contextual factors are modifiable: which have the greatest scope for change and who would benefit most; (iii) deciding on mechanisms of change(3).

Following informal discussions with respiratory experts and considering responses to Asthma + Lung UK (A+LUK) surveys of people with respiratory disease(4, 5), the authors proposed an initial model containing five domains of concern: avoidable risks that cause COPD, diagnostic delay, inadequate COPD care, low status of COPD, lack of support that people living with COPD experience(6). These categories were then circulated to an expert panel in September 2022, using SurveyMonkey to canvas opinion about the validity of this structure as well as to rate the importance of key elements that should be contained in each domain and suggest additional ones. Expert input through this process led to an increase in the emphasis on early life and transgenerational factors as well as the impact of racism.

The survey was also sent to members of an A+LUK patient group who ranked elements to confirm their importance and were able to offer additional possible items. Participants completing the questionnaire consented to their anonymised data being used for research purposes and ethical approval for this analysis was granted by Imperial College Research and Integrity Team (IREC; 20IC6625). Input from people with COPD led to an increase in the emphasis on the impact of stigma from the public and healthcare system as well as demands for legislation to tackle avoidable lung harms such as poverty and air pollution. Further stakeholder meetings were held to review and refine the model and evidence base.

The model and evidence base were circulated to an expert group for comments, additional items with two concluding meetings online for final approval in August 2023.

**Survey Canvassing**

We created 5 separate surveys based on each domain of structural violence, the first survey was sent to the expert panel, with the subsequent 4 surveys sent to the same panel two weeks later. We asked respondents to rank the importance of key factors within each domain using a five-point Likert scale from unimportant to very important. We then asked respondents to identify any additional key factors that we may have missed that contribute to the domain of structural violence. In addition to identifying and ranking key elements of structural violence we asked respondents to provide any mitigation strategies they believe would or should be implemented to target these elements of structural violence.

Once we collected responses from the expert panel, we created one large survey that contained all 5 domains of structural violence factors and sent it too COPD patients using Asthma+Lung UKs supporter base. The survey was the same format as above, using a Likert scale to rate importance and free text responses to provide mitigation strategy suggestions.

**Extended survey results**

**Expert panel**

A total of 101 people responded to the 4 surveys; the majority (68%) responded to the first survey, 10% to the second and third surveys, and 6% to the fourth and final survey.

**Table E2. Expert panel demographics**

|  | **N=101** |
| --- | --- |
| **Gender**  **Female**  **Male**  **Undisclosed** | 39 (38%)  34 (33%)  28 (29%) |
| **Age**  **18-29**  **30-44**  **45-59**  **60-79**  **80+**  **Undisclosed** | 4 (4%)  39 (39%)  19 (19%)  11 (10%)  1 (1%)  27 (27%) |
| **Ethnicity**  **White/White British**  **Mixed Race**  **Asian/ Asian British**  **Another race**  **Undisclosed** | 31 (30%)  4 (4%)  8 (8%)  8 (8%)  50 (50%) |
| **Profession**  **Nurse**  **Clinical Academic**  **Dietician**  **Doctor**  **Other**  **Physiotherapist**  **Speech and Language Therapist**  **Undisclosed** | 12 (12%)  27 (27%)  2 (2%)  25 (25%)  6 (6%)  12 (11%)  1 (1%)  16 (16%) |

**Figure E1. Expert panel- Likert rating of factors that contribute to avoidable lung harms**

**Figure E2. Expert panel- Likert rating of factors that contribute to diagnostic delay.**

Avoidable lung harms

n

|  | Very Important | Important | Moderately Important | Slightly Important | Unimportant |
| --- | --- | --- | --- | --- | --- |
| Exposures in utero (parental smoking, maternal stress) | 20 | 16 | 11 | 8 | 3 |
| Childhood exposure to passive smoke | 23 | 22 | 9 | 3 | 1 |
| Childhood exposure to indoor air pollution | 19 | 21 | 14 | 3 | 1 |
| Childhood exposure to outdoor air pollution | 17 | 20 | 15 | 5 | 1 |
| Poor nutrition in childhood | 16 | 17 | 15 | 9 | 1 |
| Childhood infections | 14 | 22 | 10 | 10 | 2 |
| Adverse childhood experiences (trauma/stress/abuse) | 7 | 14 | 14 | 17 | 6 |
| Poor housing | 16 | 24 | 12 | 5 | 1 |
| Poor access to healthcare in childhood | 17 | 18 | 18 | 4 | 1 |
| Occupational exposures (jobs with dust, fumes or chemicals) | 31 | 16 | 18 | 2 | 5 |
| Indoor air polluton in adult life | 21 | 24 | 8 | 4 | 1 |
| Outdoor air pollution in adult life | 20 | 22 | 11 | 4 | 1 |
| Lack of access to green space | 11 | 18 | 10 | 17 | 2 |
| Smoking | 51 | 5 | 1 | 0 | 1 |

Diagnostic Delay

n

|  | Very Important | Important | Moderately Important | Slightly Important | Unimportant |
| --- | --- | --- | --- | --- | --- |
| Lack of systematic approach to breathlessness | 1 | 5 | 4 | 1 | 0 |
| Symptoms normalised (symptoms such as cough and sputum linked to common cold/ viruses "smoker's cough') | 3 | 5 | 3 | 0 | 0 |
| Failure to diagnose high risk children | 1 | 5 | 3 | 0 | 2 |
| Stigma from healthcare system and self-stigmatising | 2 | 4 | 4 | 1 | 0 |
| Patients feeling unworthy of care so not accessing healthcare | 1 | 4 | 4 | 2 | 0 |
| Lower expectations of healthcare | 2 | 3 | 4 | 2 | 0 |
| Multimorbidity (having more than 1 condition) - symptoms attributed to another condition | 1 | 5 | 3 | 2 | 0 |
| People at risk for COPD have poor access to healthcare | 2 | 4 | 3 | 2 | 0 |
| Bias in health information measurements (under-researched women, people with dark skin and oximetry) | 1 | 5 | 4 | 1 | 0 |
| Lack of healthcare system incentives (targets/funding lack of governmental support) | 2 | 3 | 3 | 2 | 0 |
| Healthcare worker engagement (No cure and lack of novel treatments reducing interest from health services and healthcare workers) | 2 | 3 | 5 | 1 | 0 |
| Low public awareness of respiratory diseases | 3 | 4 | 3 | 1 | 0 |
| Lung Function measurements seen as ‘too hard’ by health services | 2 | 4 | 3 | 2 | 0 |

**Figure E3. Expert panel- Likert rating of factors that contribute to inadequate care.**

**Figure E4. Expert panel- Likert rating of factors that contribute to Low status of COPD**

Inadequate Care

n

|  | Very Important | Important | Moderately Important | Slightly Important | Unimportant |
| --- | --- | --- | --- | --- | --- |
| Poor provision of smoking cessation clinics in the community | 4 | 4 | 2 | 0 | 0 |
| Poor provision of pulmonary rehabilitation (long wait lists) | 7 | 2 | 1 | 0 | 0 |
| Self management plans not implemented or being supported by healthcare services | 4 | 2 | 1 | 3 | 0 |
| Poor provision of vaccinations | 0 | 4 | 4 | 1 | 1 |
| COPD related stigma, often due to the link between the condition and smoking | 1 | 5 | 1 | 3 | 0 |
| Clinician awareness of COPD management | 7 | 2 | 1 | 0 | 0 |
| People with COPD not aware of what care they should be receiving | 4 | 4 | 2 | 0 | 0 |
| Lack of clinical staff time | 4 | 2 | 3 | 0 | 1 |
| Social status of people with COPD | 3 | 6 | 1 | 0 | 0 |
| Lack of clinical investigations to identify and treat particular symptoms/ conditions in COPD (Bronchiectasis, Alpha-1 deficiency, sleep disordered breathing). | 4 | 2 | 1 | 3 | 0 |
| Poor access to palliative care | 4 | 2 | 2 | 1 | 1 |
| Acute COPD exacerbation (flare up) pathways are poorly supported | 4 | 5 | 0 | 1 | 0 |
| Risk of other chronic diseases is unaddressed | 4 | 3 | 1 | 1 | 1 |
| Access to peer support groups (breathe easy groups) is limited | 3 | 2 | 4 | 1 | 0 |
| Guidelines not implemented adequately | 2 | 3 | 4 | 1 | 0 |

**Figure E5. Expert panel- Likert rating of factors that contribute to lack of support for COPD patients.**

Low Status of COPD

n

|  | Very Important | Important | Moderately Important | Slightly Important | Unimportant |
| --- | --- | --- | --- | --- | --- |
| Inadequate research funding | 3 | 2 | 0 | 0 | 0 |
| Historical lack of clinical priority by health services meaning fewer treatment successes | 2 | 1 | 2 | 0 | 0 |
| limited advancement in therapies (Poor investment/ interest from government, health services and pharma in exploring new therapies) | 2 | 3 | 0 | 0 | 0 |
| Low priority for commissioners of care | 1 | 3 | 1 | 0 | 0 |
| Medical care is often only focused on exacerbations (flare-ups) in terms of reductions in healthcare costs, not quality-of-life terms or lived experience. | 3 | 1 | 1 | 0 | 0 |
| Pharmacological approaches (medication) for treating COPD is prioritised over non-pharmacological (Pulmonary rehab) treatments. | 2 | 1 | 2 | 0 | 0 |
| Stigma due to link with tobacco smoking (Society and healthcare workers viewing smoking as a behaviour choice, not an addiction). | 2 | 2 | 0 | 1 | 0 |
| Patients with COPD having low expectations of care | 2 | 3 | 0 | 0 | 0 |
| Patients lacking social and cultural capital (relationships and assets that can allow upward social mobility) | 1 | 2 | 1 | 1 | 0 |
| Low general awareness of COPD | 1 | 2 | 2 | 1 | 0 |
| A lot of care delivered by non-specialists | 3 | 1 | 1 | 0 | 0 |

Lack of Support

n

|  | Very Important | Important | Moderately Important | Slightly Important | Unimportant |
| --- | --- | --- | --- | --- | --- |
| Social isolation due to poor transport | 2 | 2 | 1 | 0 | 0 |
| Social isolation due to poor provision of local services (e.g. community centres) | 3 | 1 | 1 | 0 | 0 |
| Social isolation due to lack of recreation opportunities | 2 | 1 | 2 | 0 | 0 |
| Inability to continue work | 3 | 2 | 0 | 0 | 0 |
| Lack of social security (income) | 3 | 1 | 1 | 0 | 0 |
| Poor housing | 3 | 1 | 1 | 0 | 0 |
| Digital exclusion from online services | 4 | 0 | 0 | 1 | 0 |
| Poor access to psychological support therapies | 3 | 2 | 0 | 0 | 0 |
| Lack of peer support networks | 3 | 1 | 1 | 0 | 0 |
| Lack of public understanding | 5 | 0 | 0 | 0 | 0 |
| Anxiety about catching infections | 3 | 1 | 1 | 0 | 0 |

**Extended survey results**

**Asthma+ Lung UK supporter base**

A total of 28 people responded to the survey, including people with a COPD diagnosis (n=15), carers of someone with COPD (n=5) or people who work within respiratory medicine or advocacy (n=6). Please see table E3 for full participant demographics

**Table E3. Asthma+Lung UK supporter base demographics**

|  | **(N=28)** |
| --- | --- |
| **Gender**  **Female**  **Male** | 18 (60%)  10 (40%) |
| **Age** | 64.4 ± 14.3 |
| **Ethnicity**  **White/White British**  **Mixed Race** | 27 (96%)  1 (4%) |
| **Profession (If indicated work within respiratory medicine or advocacy)**  **Healthcare charity role**  **Academic (non-clinical)**  **Psychologist**  **Nurse**  **Clinical Academic** | 1 (4%)  1 (4%)  1 (4%)  1 (4%)  2 (6%) |

**Figure E6. Asthma+ Lung UK supporter base- Likert rating of factors that contribute to Avoidable lung harms.**

Avoidable lung harms

n

|  | Very Important | Important | Moderatley Important | Slightly Important | Unimportant |
| --- | --- | --- | --- | --- | --- |
| Exposures in utero (parental smoking, maternal stress) | 10 | 8 | 2 | 5 | 1 |
| Childhood exposure to passive smoke | 19 | 4 | 2 | 1 | 0 |
| Childhood exposure to indoor air pollution | 14 | 11 | 1 | 0 | 0 |
| Childhood exposure to outdoor air pollution | 14 | 8 | 4 | 0 | 0 |
| Poor nutrition in childhood | 8 | 9 | 6 | 3 | 0 |
| Childhood infections | 12 | 5 | 7 | 2 | 0 |
| Adverse childhood experiences (trauma/stress/abuse) | 4 | 11 | 3 | 4 | 4 |
| Poor housing | 9 | 14 | 2 | 1 | 0 |
| Poor access to healthcare in childhood | 9 | 9 | 5 | 3 | 0 |
| Occupational exposures (jobs with dust, fumes or chemicals) | 24 | 2 | 0 | 0 | 0 |
| Indoor air polluton in adult life | 16 | 10 | 0 | 0 | 0 |
| Outdoor air pollution in adult life | 12 | 12 | 2 | 0 | 0 |
| Lack of access to green space | 9 | 9 | 7 | 1 | 0 |
| Smoking | 23 | 3 | 0 | 0 | 0 |

**Figure E7. Asthma+ Lung UK supporter base- Likert rating of factors that contribute to Diagnostic delay**

Diagnostic Delay

n

|  | Very Important | Important | Moderatley Important | Slightly Important | Unimportant |
| --- | --- | --- | --- | --- | --- |
| Lack of systematic approach to breathlessness | 14 | 5 | 2 | 0 | 0 |
| Symptoms normalised (symptoms such as cough and sputum linked to common cold/ viruses "smoker's cough') | 11 | 6 | 4 | 0 | 0 |
| Failure to diagnose high risk children | 10 | 9 | 2 | 0 | 0 |
| Stigma from healthcare system and self-stigmatising | 9 | 5 | 4 | 2 | 1 |
| Patients feeling unworthy of care so not accessing healthcare | 10 | 6 | 2 | 3 | 0 |
| Lower expectations of healthcare | 10 | 5 | 4 | 2 | 0 |
| Multimorbidity (having more than 1 condition) - symptoms attributed to another condition | 9 | 9 | 3 | 0 | 0 |
| People at risk for COPD have poor access to healthcare | 9 | 7 | 3 | 1 | 1 |
| Bias in health information measurements (under-researched women, people with dark skin and oximetry) | 8 | 8 | 3 | 1 | 1 |
| Healthcare worker engagement (No cure and lack of novel treatments reducing interest from health services and healthcare workers) | 8 | 8 | 5 | 0 | 0 |
| Low public awareness of respiratory diseases | 17 | 2 | 1 | 1 | 0 |
| Lung Function measurements seen as ‘too hard’ by health services | 11 | 4 | 2 | 3 | 0 |

**Figure E8. Asthma+ Lung UK supporter base- Likert rating of factors that contribute to Inadequate care**

Inadequate Care

n

|  | Very Important | Important | Moderatley Important | Slightly Important | Unimportant |
| --- | --- | --- | --- | --- | --- |
| Poor provision of smoking cessation clinics in the community | 11 | 5 | 3 | 2 | 0 |
| Poor provision of pulmonary rehabilitation (long wait lists) | 15 | 5 | 0 | 1 | 0 |
| Self management plans not implemented or being supported by healthcare services | 14 | 6 | 0 | 0 | 1 |
| Poor provision of vaccinations | 8 | 5 | 5 | 3 | 0 |
| COPD related stigma, often due to the link between the condition and smoking | 12 | 6 | 2 | 1 | 0 |
| Clinician awareness of COPD management | 14 | 6 | 1 | 0 | 0 |
| People with COPD not aware of what care they should be receiving | 16 | 3 | 2 | 0 | 0 |
| Lack of clinical staff time | 15 | 5 | 1 | 0 | 0 |
| Social status of people with COPD | 12 | 6 | 2 | 0 | 1 |
| Lack of clinical investigations to identify and treat particular symptoms/ conditions in COPD (Bronchiectasis, Alpha-1 deficiency, sleep disordered breathing). | 16 | 6 | 0 | 1 | 0 |
| Poor access to palliative care | 9 | 10 | 1 | 1 | 0 |
| Acute COPD exacerbation (flare up) pathways are poorly supported | 15 | 5 | 0 | 1 | 0 |
| Risk of other chronic diseases is unaddressed | 11 | 6 | 3 | 1 | 0 |
| Access to peer support groups (breathe easy groups) is limited | 9 | 9 | 2 | 0 | 1 |
| Guidelines not implemented adequately | 13 | 5 | 1 | 2 | 0 |

**Figure E9. Asthma+ Lung UK supporter base- Likert rating of factors that contribute to Low status of COPD**

Low Status of COPD

|  | Very Important | Important | Moderatley Important | Slightly Important | Unimportant |
| --- | --- | --- | --- | --- | --- |
| Inadequate research funding | 13 | 7 | 1 | 0 | 0 |
| Historical lack of clinical priority by health services meaning fewer treatment successes | 11 | 7 | 1 | 0 | 0 |
| limited advancement in therapies (Poor investment/ interest from government, health services and pharma in exploring new therapies) | 12 | 7 | 2 | 0 | 0 |
| Low priority for commissioners of care | 9 | 8 | 2 | 1 | 0 |
| Medical care is often only focused on exacerbations (flare-ups) in terms of reductions in healthcare costs, not quality-of-life terms or lived experience. | 15 | 4 | 2 | 0 | 0 |
| Pharmacological approaches (medication) for treating COPD is prioritised over non-pharmacological (Pulmonary rehab) treatments. | 12 | 5 | 3 | 1 | 0 |
| Stigma due to link with tobacco smoking (Society and healthcare workers viewing smoking as a behaviour choice, not an addiction). | 11 | 5 | 5 | 0 | 0 |
| Patients with COPD having low expectations of care | 8 | 10 | 2 | 0 | 0 |
| Some patients may be lacking social and cultural capital (relationships and assets that can allow upward social mobility) | 11 | 8 | 1 | 0 | 1 |
| Low general awareness of COPD | 11 | 7 | 1 | 1 | 0 |
| A lot of care delivered by non-specialists | 7 | 11 | 2 | 1 | 0 |

**Figure E10. Asthma+ Lung UK supporter base- Likert rating of factors that contribute to Lack of support for living with COPD**

Lack of Support

n

|  | Very Important | Important | Moderatley Important | Slightly Important | Unimportant |
| --- | --- | --- | --- | --- | --- |
| Social isolation due to poor transport | 7 | 7 | 4 | 1 | 0 |
| Social isolation due to poor provision of local services (e.g. community centres) | 9 | 9 | 1 | 0 | 0 |
| Social isolation due to lack of recreation opportunities | 8 | 9 | 2 | 0 | 0 |
| Inability to continue work | 11 | 8 | 0 | 0 | 0 |
| Lack of social security (income) | 12 | 7 | 0 | 0 | 0 |
| Poor housing | 13 | 5 | 1 | 0 | 0 |
| Digital exclusion from online services | 7 | 9 | 3 | 0 | 0 |
| Poor access to psychological support therapies | 10 | 7 | 2 | 0 | 0 |
| Lack of peer support networks | 6 | 11 | 2 | 0 | 0 |
| Lack of public understanding | 8 | 8 | 2 | 0 | 0 |
| Anxiety about catching infections | 8 | 9 | 2 | 0 | 0 |

**RESULTS**

**Mitigation strategies**

We conducted an analysis on free text data from both the expert panel responses and A+LUK supporter base responses on their ideas and suggestions for mitigation strategies. The responses were read to get an overall sense of the data, the data was then organized into codes and finally into 5 main categories which best describe the overall theme of the mitigation strategy. These 4 categories include 1) Policy Makers, 2) The Public, 3) People with COPD and 4) Healthcare professionals as individuals. Table E4 below contains individual quotations.

**Table E4. Mitigation categories and quotations**

| **Category** | **Quotation** |
| --- | --- |
| Policy Makers | “Address Poverty”  “Universal basic income for all”  “Address societal factors such as structural racism and poverty”.  “Expand ULEZ” [ultra low emission zone]  “Better funding for healthcare from government” |
| The Public | “Mass education of the public”  “Better understanding from the public as to what COPD is”.  “Mass media campaigns”  “Not enough people 'jumping' up and down about it. No one in the public eye kicking up a storm”  “More awareness of symptoms. Should be more televised adverts similar to other health issues” |
| People with COPD | “Lack of public figure with condition to bring awareness to it Chronic condition with no "quick fix" so less incentive to invest time/ money in management or finding treatments”. |
| Healthcare professionals as individuals | “Simplistic questions by health care professional - e.g. Do you smoke? - No - so not COPD. Not understanding enough about what work might entail, this can't judge the risk. Health care professionals not understanding what poverty is really like”?  “People with COPD would greatly be helped with ‘Talking Therapy’”.  “For healthcare professionals not just assume you’ve done this to yourself because you have smoked” |

**Structural factors that cause and worsen COPD – final consensus.**

The final model included the following five domains, each consisting of two sub-domains

**Avoidable lung harms**: (i) Processes that impact on lung development (ii) Processes which disadvantage lung health in particular groups across the life course.

**Diagnostic Delay**: comprising (i) Healthcare factors and (ii) Norms and attitudes that mean that COPD is not diagnosed in a timely way, denying people with COPD effective treatment to improve/maintain quality of life and improve prognosis.

**Inadequate COPD Care**: ways in which the provision of care for people with COPD falls short of what is needed to ensure that they are able to enjoy the best possible health, considered as (i) Healthcare resource allocation (ii) Norms and attitudes influencing clinical practice.

**Low status of COPD:** ways in which both COPD as a condition and people with COPD are held in less regard and considered less of a priority than for other health problems (i) Institutional factors (ii) Norms and attitudes.

**Lack of Support**: factors that make living with COPD more difficult than it should be. These are categorised as (i) Socioenvironmental factors (ii) Factors that promote social isolation.

.

**Table E5: Structural violence: examples mechanisms and mitigations**

**(1) Avoidable lung harms**

Ways in which societal factors increase the risk of developing COPD in certain populations across the life course. Mechanisms and mitigations

| **Mechanism** | Reference | **Mitigation** | Reference |
| --- | --- | --- | --- |
| **Lung development**   - **Maternal health/ in utero exposures and transgenerational effects** | Bush A. Impact of early life exposures on respiratory disease. Paediatric respiratory reviews. 2021 Dec 1;40:24-32.[ <https://doi.org/10.1016/j.prrv.2021.05.006> ] (1)  Gibbs K, Collaco JM, McGrath-Morrow SA. Impact of tobacco smoke and nicotine exposure on lung development. Chest. 2016 Feb 1;149(2):552-61. [<https://doi.org/10.1378/chest.15-1858>](2)  Magnus MC, Håberg SE, Karlstad Ø, Nafstad P, London SJ, Nystad W. Grandmother's smoking when pregnant with the mother and asthma in the grandchild: the Norwegian Mother and Child Cohort Study. Thorax. 2015 Mar 1;70(3):237-43. [<http://dx.doi.org/10.1136/thoraxjnl-2014-206438>] (3)  He H, He MM, Wang H, Qiu W, Liu L, Long L, Shen Q, Zhang S, Qin S, Lu Z, Cai Y. In Utero and Childhood/Adolescence Exposure to Tobacco Smoke, Genetic Risk, and Lung Cancer Incidence and Mortality in Adulthood. American journal of respiratory and critical care medicine. 2023 Jan 15;207(2):173-82. <https://doi.org/10.1164/rccm.202112-2758OC> (4) | **Poverty reduction.**  **Tobacco control esp around conception and pregnancy**  **Increase healthcare provision.**  **Education (Sure start/ health visitors)**  **Mass media** | Gibson M, Hearty W, Craig P. The public health effects of interventions similar to basic income: a scoping review. The Lancet Public Health. 2020 Mar 1;5(3):e165-76.I:[<https://doi.org/10.1016/S2468-2667(20)30005-0>] (5)  Levy DT, Yuan Z, Luo Y, Mays D. Seven years of progress in tobacco control: an evaluation of the effect of nations meeting the highest level MPOWER measures between 2007 and 2014. Tobacco control. 2018 Jan 1;27(1):50-7.  [<http://dx.doi.org/10.1136/tobaccocontrol-2016-053381>](6)  Melhuish E, Belsky J, Barnes J. Evaluation and value of Sure Start. Archives of disease in childhood. 2010 Mar 1;95(3):159-61. [<http://dx.doi.org/10.1136/adc.2009.161018>] (7)  Wakefield MA, Loken B, Hornik RC. Use of mass media campaigns to change health behaviour. The lancet. 2010 Oct 9;376(9748):1261-71. [<https://doi.org/10.1016/S0140-6736(10)60809-4>] (8) |
| **Lung health disadvantage childhood onwards**   - **Passive smoking** - **Risk of smoking uptake** - **Respiratory infections** - **Poverty** - **Poor housing** - **Outdoor/indoor air pollution** | Leonardi-Bee J, Jere ML, Britton J. Exposure to parental and sibling smoking and the risk of smoking uptake in childhood and adolescence: a systematic review and meta-analysis. Thorax. 2011 Oct 1;66(10):847-55.[ <http://dx.doi.org/10.1136/thx.2010.153379>] (9)  Vrinten C, Parnham JC, Filippidis FT, Hopkinson NS, Laverty AA. Risk factors for adolescent smoking uptake: Analysis of prospective data from the UK Millennium Cohort Study. Tobacco Induced Diseases. 2022;20.[ doi: 10.18332/tid/152321](10)  Laverty AA, Filippidis FT, Taylor-Robinson D, Millett C, Bush A, Hopkinson NS. Smoking uptake in UK children: analysis of the UK Millennium Cohort Study. Thorax. 2019 Jun 1;74(6):607-10. [[10.1136/thoraxjnl-2018-212254](https://doi.org/10.1136/thoraxjnl-2018-212254)] (11)  Patel JH, Amaral AF, Minelli C, Elfadaly FG, Mortimer K, El Sony A, El Rhazi K, Seemungal TA, Mahesh PA, Obaseki DO, Denguezli M. Chronic airflow obstruction attributable to poverty in the multinational Burden of Obstructive Lung Disease (BOLD) study. Thorax. 2023 Jul 7. [<http://dx.doi.org/10.1136/thorax-2022-218668>](12)  Burney P. Chronic respiratory disease–the acceptable epidemic?. Clinical Medicine. 2017 Feb;17(1):29 [[10.7861/clinmedicine.17-1-29](https://doi.org/10.7861%2Fclinmedicine.17-1-29)](13)  Balogun B, Rankl F, Wilson W. Health inequalities cold or damp homes. House of Commons Library. 2023 Feb 16 [https://researchbriefings.files.parliament.uk/documents/CBP-9696/CBP-9696.pdf] (14)  Wimalasena NN, Chang-Richards A, Wang KI, Dirks KN. Housing risk factors associated with respiratory disease: a systematic review. International journal of environmental research and public health. 2021 Mar 10;18(6):2815. [[10.3390/ijerph18062815](https://doi.org/10.3390%2Fijerph18062815)] (15)  Allinson JP, Chaturvedi N, Wong A, Shah I, Donaldson GC, Wedzicha JA, Hardy R. Early childhood lower respiratory tract infection and premature adult death from respiratory disease in Great Britain: a national birth cohort study. The Lancet. 2023 Apr 8;401(10383):1183-93. <https://doi.org/10.1016/S0140-6736(23)00131-9> (16)  Duan P, Wang Y, Lin R, Zeng Y, Chen C, Yang L, Yue M, Zhong S, Wang Y, Zhang Q. Impact of early life exposures on COPD in adulthood: A systematic review and meta‐analysis. Respirology. 2021 Dec;26(12):1131-51. [https://doi.org/10.1111/resp.1414**4**](https://doi.org/10.1111/resp.14144) (17) | **Smoking cessation**  **Reductions in poverty**  **Social security**  **Education**  **Legislation**  **Low emission zones**  **Clean air law**  **Active travel/ town planning /schools streets**  **Air filtration**  **UBI**  **Sure start/ health visitors** | Bo Y, Chang LY, Guo C, Lin C, Lau AK, Tam T, Lao XQ. Reduced ambient PM2. 5, better lung function, and decreased risk of chronic obstructive pulmonary disease. Environment International. 2021 Nov 1;156:106706. [<https://doi.org/10.1016/j.envint.2021.106706>] (18)  Mayor of London. Air Quality Monitoring Study: London School Streets. London Assembly. 2021 Mar.[ <https://www.london.gov.uk/sites/default/files/school_streets_monitoring_study_march21.pdf>] (19)  Gibson M, Hearty W, Craig P. The public health effects of interventions similar to basic income: a scoping review. The Lancet Public Health. 2020 Mar 1;5(3):e165-76. [<https://doi.org/10.1016/S2468-2667(20)30005-0>]  (5)  Mudway, I.S., Dundas, I., Wood, H.E., Marlin, N., Jamaludin, J.B., Bremner, S.A., Cross, L., Grieve, A., Nanzer, A., Barratt, B.M. and Beevers, S., 2019. Impact of London's low emission zone on air quality and children's respiratory health: a sequential annual cross-sectional study. *The Lancet Public Health*, *4*(1), pp.e28-e40.[ <https://doi.org/10.1016/S2468-2667(18)30202-0>] (20)  Chamberlain RC, Fecht D, Davies B, Laverty AA. Health effects of low emission and congestion charging zones: a systematic review. The Lancet Public Health. 2023 Jul 1;8(7):e559-74.[ <https://doi.org/10.1016/S2468-2667(23)00120-2>] (21)  Deming D. Early childhood intervention and life-cycle skill development: Evidence from Head Start. American Economic Journal: Applied Economics. 2009 Jul 1;1(3):111-34. DOI: 10.1257/app.1.3.111(22) |
| **Lung health disadvantage childhood onwards**  **-Adverse childhood experiences** | Suglia SF, Ryan L, Laden F, Dockery D, Wright RJ. Violence exposure, a chronic psychosocial stressor, and childhood lung function. Psychosomatic medicine. 2008 Feb;70(2):160. [10.1097/PSY.0b013e318160687c](https://doi.org/10.1097%2FPSY.0b013e318160687c) (23)  Bailey BA. Partner violence during pregnancy: prevalence, effects, screening, and management. International journal of women's health. 2010 Aug 9:183-97 [10.2147/ijwh.s8632](https://doi.org/10.2147%2Fijwh.s8632) (24) | **Education**  **Reduce poverty.**  **Increased access to CAMHS** | Gibson M, Hearty W, Craig P. The public health effects of interventions similar to basic income: a scoping review. The Lancet Public Health. 2020 Mar 1;5(3):e165-76. [:<https://doi.org/10.1016/S2468-2667(20)30005-0>] |
| **Lung health disadvantage childhood onwards**  **-Poor nutrition (Obesity/ malnutrition)** | van Abeelen AF, Elias SG, de Jong PA, Grobbee DE, Bossuyt PM, van der Schouw YT, Roseboom TJ, Uiterwaal CS. Famine in the young and risk of later hospitalization for COPD and asthma. PLoS One. 2013 Dec 23;8(12):e82636. <https://doi.org/10.1371/journal.pone.0082636>  (25)  Yang IA, Jenkins CR, Salvi SS. Chronic obstructive pulmonary disease in never-smokers: risk factors, pathogenesis, and implications for prevention and treatment. The Lancet Respiratory Medicine. 2022 Apr 12.:<https://doi.org/10.1016/S2213-2600(21)00506-3> (26)  van Iersel LE, Beijers RJ, Gosker HR, Schols AM. Nutrition as a modifiable factor in the onset and progression of pulmonary function impairment in COPD: A systematic review. Nutrition reviews. 2022 Jun 1;80(6):1434-44. [<https://doi.org/10.1093/nutrit/nuab077>](27)  Varraso R, Chiuve SE, Fung TT, Barr RG, Hu FB, Willett WC, Camargo CA. Alternate Healthy Eating Index 2010 and risk of chronic obstructive pulmonary disease among US women and men: prospective study. bmj. 2015 Feb 3;350. [<https://doi.org/10.1136/bmj.h286>](28)  Varraso R, Dumas O, Boggs KM, Willett WC, Speizer FE, Camargo CA. Processed meat intake and risk of chronic obstructive pulmonary disease among middle-aged women. EClinicalMedicine. 2019 Sep 1;14:88-95. [https://doi.org/10.1016/j.eclinm.2019.07.014](29) | **Policy changes (cheaper healthier food)**  **Town planning**  **Education**  **Free school meals for all**  **Reductions in poverty/UBI**  **Sure, start centres/ health visitors.**  **Media campaigns**  **Legislation**  **Social security** | Gibson M, Hearty W, Craig P. The public health effects of interventions similar to basic income: a scoping review. The Lancet Public Health. 2020 Mar 1;5(3):e165-76. [<https://doi.org/10.1016/S2468-2667(20)30005-0>] (5)  Vik FN, Van Lippevelde W, Øverby NC. Free school meals as an approach to reduce health inequalities among 10–12-year-old Norwegian children. BMC Public Health. 2019 Dec;19(1):1-8.  https://doi.org/10.1186/s12889-019-7286-z  (30) |
| **Lung health disadvantage childhood onwards**  **-Systemic racism within and between countries** | Balmes JR. Place Matters: Residential Racial Segregation and Chronic Obstructive Pulmonary Disease. American Journal of Respiratory and Critical Care Medicine. 2021 Sep 1;204(5):496-8. <https://doi.org/10.1164/rccm.202009-3721OC>    (31)  Mamary AJ, Stewart JI, Kinney GL, Hokanson JE, Shenoy K, Dransfield MT, Foreman MG, Vance GB, Criner GJ, COPDGene® Investigators. Race and gender disparities are evident in COPD underdiagnoses across all severities of measured airflow obstruction. Chronic Obstructive Pulmonary Diseases: Journal of the COPD Foundation. 2018;5(3):177. [ [10.15326/jcopdf.5.3.2017.0145](https://doi.org/10.15326%2Fjcopdf.5.3.2017.0145)] (32)  Burney P. Chronic respiratory disease–the acceptable epidemic?. Clinical Medicine. 2017 Feb;17(1):29 [[10.7861/clinmedicine.17-1-29](https://doi.org/10.7861%2Fclinmedicine.17-1-29)](13) | **Legislation**  **Reparations**  **Social-cultural change**  **Education**  **Medical Education** | Bailey ZD, Krieger N, Agénor M, Graves J, Linos N, Bassett MT. Structural racism and health inequities in the USA: evidence and interventions. The lancet. 2017 Apr 8;389(10077):1453-63. <https://doi.org/10.1016/S0140-6736(17)30569-X> (33) |
| **Lung health disadvantage childhood onwards**  **-Reduced access to healthcare** | Yang IA, Jenkins CR, Salvi SS. Chronic obstructive pulmonary disease in never-smokers: risk factors, pathogenesis, and implications for prevention and treatment. The Lancet Respiratory Medicine. 2022 Apr 12. <https://doi.org/10.1016/S2213-2600(21)00506-3> (26) | **Easy access clinics in the community**  **Link with schooling/ school nurses**  **Employment rights**  **Political action**  **Reduce poverty.**  **Invest in family/ community health.**  **Social reform**  **Public health policies** | Yu SW, Hill C, Ricks ML, Bennet J, Oriol NE. The scope and impact of mobile health clinics in the United States: a literature review. International journal for equity in health. 2017 Dec;16(1):1-2.  https://doi.org/10.1186/s12939-017-0671-2  (34) |
| **Lung health disadvantage childhood onwards**  **-Occupational exposures** | Yang IA, Jenkins CR, Salvi SS. Chronic obstructive pulmonary disease in never-smokers: risk factors, pathogenesis, and implications for prevention and treatment. The Lancet Respiratory Medicine. 2022 Apr 12.:<https://doi.org/10.1016/S2213-2600(21)00506-3> (26)  Murgia N, Gambelunghe A. Occupational COPD—The most under‐recognized occupational lung disease?. Respirology. 2022 Jun;27(6):399-410.  [10.1111/resp.14272](https://doi.org/10.1111/resp.14272) (35) | **Active lung health**  **Workplace legislation**  **PPE – provision and enforcement**  **Laws to protect workers**  **Organization.**  **Education** | Robson LS, Clarke JA, Cullen K, Bielecky A, Severin C, Bigelow PL, Irvin E, Culyer A, Mahood Q. The effectiveness of occupational health and safety management system interventions: a systematic review. Safety science. 2007 Mar 1;45(3):329-53. <https://doi.org/10.1016/j.ssci.2006.07.003> (36)  Malinowski B, Minkler M, Stock L. Labor unions: a public health institution. American journal of public health. 2015 Feb;105(2):261-71.[ [10.2105/AJPH.2014.302309](https://doi.org/10.2105%2FAJPH.2014.302309)](37) |
| **Lung health disadvantage childhood onwards**  **-Environmental degradation/ pollution/ climate disruption** | Yang IA, Jenkins CR, Salvi SS. Chronic obstructive pulmonary disease in never-smokers: risk factors, pathogenesis, and implications for prevention and treatment. The Lancet Respiratory Medicine. 2022 Apr 12<https://doi.org/10.1016/S2213-2600(21)00506-3> (26)  Sarkar C, Zhang B, Ni M, Kumari S, Bauermeister S, Gallacher J, Webster C. Environmental correlates of chronic obstructive pulmonary disease in 96 779 participants from the UK Biobank: a cross-sectional, observational study. The Lancet Planetary Health. 2019 Nov 1;3(11):e478-90. [[10.1016/S2542-5196(19)30214-1](https://doi.org/10.1016/s2542-5196(19)30214-1)] (38)  Shin S, Bai L, Burnett RT, Kwong JC, Hystad P, van Donkelaar A, Lavigne E, Weichenthal S, Copes R, Martin RV, Kopp A. Air pollution as a risk factor for incident chronic obstructive pulmonary disease and asthma. A 15-year population-based cohort study. American journal of respiratory and critical care medicine. 2021 May 1;203(9):1138-48. [[10.1164/rccm.201909-1744OC](https://doi.org/10.1164/rccm.201909-1744oc)] (39)  van Gemert F, Kirenga B, Chavannes N, Kamya M, Luzige S, Musinguzi P, Turyagaruka J, Jones R, Tsiligianni I, Williams S, de Jong C. Prevalence of chronic obstructive pulmonary disease and associated risk factors in Uganda (FRESH AIR Uganda): a prospective cross-sectional observational study. The Lancet Global Health. 2015 Jan 1;3(1):e44-51. [https://doi.org/10.1016/S2214-109X(14)70337-7].(40) | **Green policies**  **Legislation**  **ULEZ Clean air law**  **Active travel/ town planning**  **Air filtration**  **Reduce poverty.** | Bo Y, Chang LY, Guo C, Lin C, Lau AK, Tam T, Lao XQ. Reduced ambient PM2. 5, better lung function, and decreased risk of chronic obstructive pulmonary disease. Environment International. 2021 Nov 1;156:106706.[ <https://doi.org/10.1016/j.envint.2021.106706>]  (18)  Chamberlain RC, Fecht D, Davies B, Laverty AA. Health effects of low emission and congestion charging zones: a systematic review. The Lancet Public Health. 2023 Jul 1;8(7):e559-74 [<https://doi.org/10.1016/S2468-2667(23)00120-2>]. (21)  Aitsi-Selmi A, Hopkinson NS. Breathlessness, physical activity and sustainability of healthcare. European Respiratory Journal. 2015 Jan 1;45(1):284-5. [[10.1183/09031936.00112614](https://doi.org/10.1183/09031936.00112614)](41) |
| **Lung health disadvantage childhood onwards**  **-Smoking** | Royal College of Physicians. Smoking and Health. 1962 https://www.rcplondon.ac.uk/projects/outputs/smoking-and-health-1962(42) | **Polluters pay levy.**  **New Zealand model**  **Tobacco Control**  **Investment in Cessation**  **Increased local authority funding** | Fidler JA, West R. Changes in smoking prevalence in 16–17‐year‐old versus older adults following a rise in legal age of sale: findings from an English population study. Addiction. 2010 Nov;105(11):1984-8. [10.1111/j.1360-0443.2010.03039.x](https://doi.org/10.1111/j.1360-0443.2010.03039.x) (17) |
| **Lung health disadvantage childhood onwards**  **-Drug use** | Yang IA, Jenkins CR, Salvi SS. Chronic obstructive pulmonary disease in never-smokers: risk factors, pathogenesis, and implications for prevention and treatment. The Lancet Respiratory Medicine. 2022 Apr 12.<https://doi.org/10.1016/S2213-2600(21)00506-3> (26)  Burhan H, Young R, Byrne T, Peat R, Furlong J, Renwick S, Elkin T, Oelbaum S, Walker PP. Screening heroin smokers attending community drug services for COPD. Chest. 2019 Feb 1;155(2):279-87.[ <https://doi.org/10.1016/j.chest.2018.08.1049>] (43) | **Reduce poverty.**  **Increase social security**  **Investment in drug and alcohol services**  **Legislation**  **Public health investment** | Haagh L, Rohregger B. Universal basic income policies and their potential for addressing health inequities: Transformative approaches to a healthy, prosperous life for all. World Health Organization. Regional Office for Europe; 2019. WHO/EURO:2019-3533-43292-60676(44) |
| **Lung health disadvantage childhood onwards**  **-Poor physical activity** | Yang IA, Jenkins CR, Salvi SS. Chronic obstructive pulmonary disease in never-smokers: risk factors, pathogenesis, and implications for prevention and treatment. The Lancet Respiratory Medicine. 2022 Apr 12.:<https://doi.org/10.1016/S2213-2600(21)00506-3> (26)  Hopkinson NS, Polkey MI. Does physical inactivity cause chronic obstructive pulmonary disease?. Clinical science. 2010 May 1;118(9):565-72. [[10.1042/CS20090458](https://doi.org/10.1042/cs20090458)](45) | **Green policies**  **Legislation**  **ULEZ Clean air law**  **Active travel/ town planning**  **Education**  **Mass media**  **Reduce poverty** | Jennings V, Bamkole O. The relationship between social cohesion and urban green space: An avenue for health promotion. International journal of environmental research and public health. 2019 Feb;16(3):452. <https://doi.org/10.3390/ijerph16030452>  (46)  Chamberlain RC, Fecht D, Davies B, Laverty AA. Health effects of low emission and congestion charging zones: a systematic review. The Lancet Public Health. 2023 Jul 1;8(7):e559-74. <https://doi.org/10.1016/S2468-2667(23)00120-2> (21) |

**(2) Diagnostic Delay**

Ways in which COPD is not diagnosed in a timely way that would allow the most effective treatment and improve/maintain quality of life and improve prognosis. Mechanisms and mitigations

| **Mechanisms** | References | **Mitigations** | References |
| --- | --- | --- | --- |
| **Healthcare factors**   - **Lack of systematic approach to breathlessness** | Jones RC, Price D, Ryan D, Sims EJ, von Ziegenweidt J, Mascarenhas L, Burden A, Halpin DM, Winter R, Hill S, Kearney M. Opportunities to diagnose chronic obstructive pulmonary disease in routine care in the UK: a retrospective study of a clinical cohort. The Lancet Respiratory Medicine. 2014 Apr 1;2(4):267-76. <https://doi.org/10.1016/S2213-2600(14)70008-6> (47)  Elbehairy AF, Quint JK, Rogers J, Laffan M, Polkey MI, Hopkinson NS. Patterns of breathlessness and associated consulting behaviour: results of an online survey. Thorax. 2019 Aug 1;74(8):814-7.[ [10.1136/thoraxjnl-2018-212950](https://doi.org/10.1136/thoraxjnl-2018-212950)] (48)  Hopkinson NS, Baxter N, London Respiratory Network. Breathing SPACE—a practical approach to the breathless patient. NPJ Primary Care Respiratory Medicine. 2017 Jan 30;27(1):5. [[10.1038/s41533-016-0006-6](https://doi.org/10.1038/s41533-016-0006-6)](49) | **Funding**  **Mass media**  **Education** | Wakefield MA, Loken B, Hornik RC. Use of mass media campaigns to change health behaviour. The lancet. 2010 Oct 9;376(9748):1261-71. [<https://doi.org/10.1016/S0140-6736(10)60809-4>](8)  Taskforce for Lung health. Finding Lung disease Early. 2023. Available from [https://www.taskforceforlunghealth.org.uk/taskforce/plan/diagnosis] Accessed on 24/07/2023.(50) |
| **Healthcare Factors**  **-Symptoms normalised** | Doe GE, Williams MT, Chantrell S, Steiner MC, Armstrong N, Hutchinson A, Evans RA. Diagnostic delays for breathlessness in primary care: a qualitative study to investigate current care and inform future pathways. British Journal of General Practice. 2023 Jun 1;73(731):e468-77. [https://doi.org/10.3399/BJGP.2022.0475] (51) | **Education**  **Better diagnostic pathways** | Hall I, Walker S, Holgate ST. Respiratory research in the UK: investing for the next 10 years. Thorax. 2022 Sep 1;77(9):851-3. <http://dx.doi.org/10.1136/thoraxjnl-2021-218459> (52) |
| **Healthcare Factors**   - **Failure to diagnose high risk children and adults:** | Yang IA, Jenkins CR, Salvi SS. Chronic obstructive pulmonary disease in never-smokers: risk factors, pathogenesis, and implications for prevention and treatment. The Lancet Respiratory Medicine. 2022 Apr 12. <https://doi.org/10.1016/S2213-2600(21)00506-3> (26) | **Reducing poverty**  **Social care/ security investment**  **Invest in treatment pathways.**  **Links with school/ school nurses**  **Spiro in year 2**  **Screening** | Gibson M, Hearty W, Craig P. The public health effects of interventions similar to basic income: a scoping review. The Lancet Public Health. 2020 Mar 1;5(3):e165-76.[ I:<https://doi.org/10.1016/S2468-2667(20)30005-0>] (5)  Hall I, Walker S, Holgate ST. Respiratory research in the UK: investing for the next 10 years. Thorax. 2022 Sep 1;77(9):851-3. <http://dx.doi.org/10.1136/thoraxjnl-2021-218459> (52)  Burhan H, Young R, Byrne T, Peat R, Furlong J, Renwick S, Elkin T, Oelbaum S, Walker PP. Screening heroin smokers attending community drug services for COPD. Chest. 2019 Feb 1;155(2):279-87.[ https://doi.org/10.1016/j.chest.2018.08.1049] (40) |
| **Diagnostic Delay**   - **Stigma from healthcare system and self-stigmatising** | Mathioudakis AG, Ananth S, Vestbo J. Stigma: an unmet public health priority in COPD. The Lancet Respiratory Medicine. 2021 Sep 1;9(9):955-6. <https://doi.org/10.1016/S2213-2600(21)00316-7> (53)  Brighton LJ, Chilcot J, Maddocks M. Social dimensions of chronic respiratory disease: stigma, isolation, and loneliness. Current Opinion in Supportive and Palliative Care. 2022 Dec 1;16(4):195-202. <https://doi.org/10.1097/SPC.0000000000000616> (54) | **Education medicine/ nurses**  **Social-cultural change**  **Mass media** | Wakefield MA, Loken B, Hornik RC. Use of mass media campaigns to change health behaviour. The lancet. 2010 Oct 9;376(9748):1261-71.[ <https://doi.org/10.1016/S0140-6736(10)60809-4>] (4)  Nyblade L, Stockton MA, Giger K, Bond V, Ekstrand ML, Lean RM, Mitchell EM, Nelson LR, Sapag JC, Siraprapasiri T, Turan J. Stigma in health facilities: why it matters and how we can change it. BMC medicine. 2019 Dec;17:1-5. https://doi.org/10.1186/s12916-019-1256-2 (55) |
| **Diagnostic Delay**   - **Patients feeling unworthy of care** | Mathioudakis AG, Ananth S, Vestbo J. Stigma: an unmet public health priority in COPD. The Lancet Respiratory Medicine. 2021 Sep 1;9(9):955-6. <https://doi.org/10.1016/S2213-2600(21)00316-7> (53) | **New face of COPD?**  **Social-cultural change**  **Education**  **Investments in PR** | Nyblade L, Stockton MA, Giger K, Bond V, Ekstrand ML, Lean RM, Mitchell EM, Nelson LR, Sapag JC, Siraprapasiri T, Turan J. Stigma in health facilities: why it matters and how we can change it. BMC medicine. 2019 Dec;17:1-5 https://doi.org/10.1186/s12916-019-1256-2.(55) |
| **Diagnostic Delay**   - **Lower expectations of healthcare** | Asthma + Lung UK Failing on the fundamentals report https://cdn.shopify.com/s/files/1/0221/4446/files/COPD_survey.pdf(56)  Buttery SC, Zysman M, Vikjord SA, Hopkinson NS, Jenkins C, Vanfleteren LE. Contemporary perspectives in COPD: patient burden, the role of gender and trajectories of multimorbidity. Respirology. 2021 May;26(5):419-41. [<https://doi.org/10.1111/resp.14032>] (57) | **Reductions in poverty**  **Social-cultural change**  **Education** | Haagh L, Rohregger B. Universal basic income policies and their potential for addressing health inequities: Transformative approaches to a healthy, prosperous life for all. World Health Organization. Regional Office for Europe; 2019. WHO/EURO:2019-3533-43292-60676 (44) |
| **Healthcare Factors**   - **Multimorbidity accumulates** | Burke H, Wilkinson TM. Unravelling the mechanisms driving multimorbidity in COPD to develop holistic approaches to patient-centred care. European Respiratory Review. 2021 Jun 30;30(160) [10.1183/16000617.0041-2021](https://doi.org/10.1183/16000617.0041-2021) .(58) | **Investment in healthcare**  **Invest in treatment pathways**  **Guidelines** | Masters R, Anwar E, Collins B, Cookson R, Capewell S. Return on investment of public health interventions: a systematic review. J Epidemiology Community Health. 2017 Aug 1;71(8):827-34. <http://dx.doi.org/10.1136/jech-2016-208141>(59) |
| **Healthcare factors**  **-Lower access to healthcare** | Raju S, Keet CA, Paulin LM, Matsui EC, Peng RD, Hansel NN, McCormack MC. Rural residence and poverty are independent risk factors for chronic obstructive pulmonary disease in the United States. American journal of respiratory and critical care medicine. 2019 Apr 15;199(8):961-9. [10.1164/rccm.201807-1374OC](https://doi.org/10.1164%2Frccm.201807-1374OC) (60) | **Education**  **Reduction in poverty**  **Easy access clinics in the community**  **Political action**  **Reduce poverty.**  **Invest in family/ community health.**  **Social reform**  **Public health policies** | Yu SW, Hill C, Ricks ML, Bennet J, Oriol NE. The scope and impact of mobile health clinics in the United States: a literature review. International journal for equity in health. 2017 Dec;16(1):1-2. https://doi.org/10.1186/s12939-017-0671-2 (34)  Haagh L, Rohregger B. Universal basic income policies and their potential for addressing health inequities: Transformative approaches to a healthy, prosperous life for all. World Health Organization. Regional Office for Europe; 2019. WHO/EURO:2019-3533-43292-60676 (44) |
| **Norms and Attitudes**   - **Racial bias in measurements** | Elmaleh-Sachs A, Balte P, Oelsner EC, Allen NB, Baugh A, Bertoni AG, Hankinson JL, Pankow J, Post WS, Schwartz JE, Smith BM. Race/ethnicity, spirometry reference equations, and prediction of incident clinical events: the Multi-Ethnic Study of Atherosclerosis (MESA) Lung Study. American journal of respiratory and critical care medicine. 2022 Mar 15;205(6):700-10. <https://doi.org/10.1164/rccm.202107-1612OC> (61)  Jamali H, Castillo LT, Morgan CC, Coult J, Muhammad JL, Osobamiro OO, Parsons EC, Adamson R. Racial disparity in oxygen saturation measurements by pulse oximetry: evidence and implications. Annals of the American Thoracic Society. 2022 Dec;19(12):1951-64. [https://doi.org/10.1513/AnnalsATS.202203-270CME] (62) | **EDI should be part of all research.**  **Stop race correction in spiro** | Bailey ZD, Krieger N, Agénor M, Graves J, Linos N, Bassett MT. Structural racism and health inequities in the USA: evidence and interventions. The lancet. 2017 Apr 8;389(10077):1453-63. <https://doi.org/10.1016/S0140-6736(17)30569-X> (30) |
| **Healthcare factors**   - **Lack of healthcare system incentives** | Burney P. Chronic respiratory disease–the acceptable epidemic?. Clinical Medicine. 2017 Feb;17(1):29. [10.7861/clinmedicine.17-1-29](https://doi.org/10.7861%2Fclinmedicine.17-1-29)(13) | **More governmental funding**  **Funding for research** | Hall I, Walker S, Holgate ST. Respiratory research in the UK: investing for the next 10 years. Thorax. 2022 Sep 1;77(9):851-3. <http://dx.doi.org/10.1136/thoraxjnl-2021-218459> (52) |
| **Norms and attitudes**   - **Therapeutic nihilism** | Burney P. Chronic respiratory disease–the acceptable epidemic?. Clinical Medicine. 2017 Feb;17(1):29. [10.7861/clinmedicine.17-1-29](https://doi.org/10.7861%2Fclinmedicine.17-1-29) (13)  Zoumot Z, Jordan S, Hopkinson NS. Emphysema: time to say farewell to therapeutic nihilism. Thorax. 2014 Nov 1;69(11):973-5. [<http://dx.doi.org/10.1136/thoraxjnl-2014-205667>] (63) | **More governmental funding**  **Funding for research**  **Education (HCW)** | Hall I, Walker S, Holgate ST. Respiratory research in the UK: investing for the next 10 years. Thorax. 2022 Sep 1;77(9):851-3. <http://dx.doi.org/10.1136/thoraxjnl-2021-218459> (52)  Hopkinson NS. Lung volume reduction: apex treatments and the ecology of chronic obstructive pulmonary disease care. American Journal of Respiratory and Critical Care Medicine. 2019 Dec 1;200(11):1329-31. [<https://doi.org/10.1164/rccm.201908-1528ED>**] (64)**  Yousuf A, Brightling CE. Biologic drugs: a new target therapy in COPD?. COPD: Journal of Chronic Obstructive Pulmonary Disease. 2018 Mar 4;15(2):99-107.[ <https://doi.org/10.1164/rccm.201908-1528ED>**] (65)** |
| **Norms and attitudes**   - **Low public understanding of lung diseases** | Walters JA, Hansen EC, Walters EH, Wood-Baker R. Under-diagnosis of chronic obstructive pulmonary disease: a qualitative study in primary care. Respiratory medicine. 2008 May 1;102(5):738-43 [10.1016/j.rmed.2007.12.008](https://doi.org/10.1016/j.rmed.2007.12.008).(66) | **Mass media**  **Education**  **New face for COPD**  **Active public health campaigns** | Wakefield MA, Loken B, Hornik RC. Use of mass media campaigns to change health behaviour. The lancet. 2010 Oct 9;376(9748):1261-71.[ <https://doi.org/10.1016/S0140-6736(10)60809-4>] (4) |
| **Healthcare factors**  **-Lung function test provision** | Walters JA, Hansen EC, Walters EH, Wood-Baker R. Under-diagnosis of chronic obstructive pulmonary disease: a qualitative study in primary care. Respiratory medicine. 2008 May 1;102(5):738-43. [10.1016/j.rmed.2007.12.008](https://doi.org/10.1016/j.rmed.2007.12.008) (66) | **Better remuneration to practices/nurses who do these.**  **More governmental funding**  **Funding for research** | Hall I, Walker S, Holgate ST. Respiratory research in the UK: investing for the next 10 years. Thorax. 2022 Sep 1;77(9):851-3. <http://dx.doi.org/10.1136/thoraxjnl-2021-218459> (52) |

**(3) Inadequate COPD Care**

Ways in which the provision of care for people with COPD falls short of what is needed to ensure that they are able to enjoy the best possible health.

Mechanisms and mitigations

| **Mechanisms** | Refs | **Mitigations** | Ref |
| --- | --- | --- | --- |
| **Poor Provision - High value care** | Philip K, Gaduzo S, Rogers J, Laffan M, Hopkinson NS. Patient experience of COPD care: outcomes from the British lung Foundation patient Passport. BMJ open respiratory research. 2019 Sep 1;6(1):e000478. <http://dx.doi.org/10.1136/bmjresp-2019-000478> (67)  Meiwald A, Gara-Adams R, Rowlandson A, Ma Y, Watz H, Ichinose M, Scullion J, Wilkinson T, Bhutani M, Weston G, Adams EJ. Qualitative Validation of COPD Evidenced Care Pathways in Japan, Canada, England, and Germany: Common Barriers to Optimal COPD Care. International journal of chronic obstructive pulmonary disease. 2022 Jul 1:1507-21.  [10.2147/COPD.S360983](https://doi.org/10.2147/copd.s360983)(68) | **Investment in health and social care**  **Government investment into research** | Hall I, Walker S, Holgate ST. Respiratory research in the UK: investing for the next 10 years. Thorax. 2022 Sep 1;77(9):851-3. <http://dx.doi.org/10.1136/thoraxjnl-2021-218459> (52)  Masters R, Anwar E, Collins B, Cookson R, Capewell S. Return on investment of public health interventions: a systematic review. J Epidemiology Community Health. 2017 Aug 1;71(8):827-34. <http://dx.doi.org/10.1136/jech-2016-208141>(59) |
| **Norms and attitudes**   - **COPD related stigma due to historical smoking behaviour** | Woo S, Zhou W, Larson JL. Stigma experiences in people with chronic obstructive pulmonary disease: an integrative review. International Journal of Chronic Obstructive Pulmonary Disease. 2021 Jun 4:1647-59. [ [10.2147/COPD.S306874](https://doi.org/10.2147%2FCOPD.S306874)](69)  Madawala S, Osadnik CR, Warren N, Kasiviswanathan K, Barton C. Healthcare experiences of adults with Chronic Obstructive Pulmonary Disease (COPD) across community care settings: a meta-ethnography. ERJ Open Research. 2022 Jan 1.[10.1183/23120541.00581-2022] (70)  Brighton LJ, Chilcot J, Maddocks M. Social dimensions of chronic respiratory disease: stigma, isolation, and loneliness. Current Opinion in Supportive and Palliative Care. 2022 Dec 1;16(4):195-202. <https://doi.org/10.1097/SPC.0000000000000616>(54) | **Education (medical/nursing school)**  **Investment in healthcare**  **Education** | Hurst JR, Winders T, Worth H, Bhutani M, Gruffydd-Jones K, Stolz D, Dransfield MT. A patient charter for chronic obstructive pulmonary disease. Advances in therapy. 2021 Jan;38:11-23. [https://doi.org/10.1007/s12325-020-01577-7] (71) |
| **Poor provision of care**  **-PR**  **-Smoking Cessation**  **-Vaccinations**  **-Self- management support/plans**  **-Palliative care** | Philip K, Gaduzo S, Rogers J, Laffan M, Hopkinson NS. Patient experience of COPD care: outcomes from the British lung Foundation patient Passport. BMJ open respiratory research. 2019 Sep 1;6(1):e000478 <http://dx.doi.org/10.1136/bmjresp-2019-000478>.(67)  Halpin DM. Palliative care for people with COPD: effective but underused. European Respiratory Journal. 2018 Feb 1;51(2). <https://doi.org/10.1183/13993003.02645-2017> (72)  Bloom C, Slaich B, Morales DR, Smeeth L, Stone P, Quint JK. Low uptake of palliative care support for COPD patients within primary care in the UK. <https://doi.org/10.1183/13993003.01879-2017>(73) | **Investment in health and social care**  **Government investment into research**  **Local authority investment**  **Public health investment** | Hall I, Walker S, Holgate ST. Respiratory research in the UK: investing for the next 10 years. Thorax. 2022 Sep 1;77(9):851-3. <http://dx.doi.org/10.1136/thoraxjnl-2021-218459> (52)  Masters R, Anwar E, Collins B, Cookson R, Capewell S. Return on investment of public health interventions: a systematic review. J Epidemiology Community Health. 2017 Aug 1;71(8):827-34 <http://dx.doi.org/10.1136/jech-2016-208141>.(59) |
| **Poor provision of care**   - **AECOPD care pathways poor** | Philip K, Gaduzo S, Rogers J, Laffan M, Hopkinson NS. Patient experience of COPD care: outcomes from the British lung Foundation patient Passport. BMJ open respiratory research. 2019 Sep 1;6(1):e000478 <http://dx.doi.org/10.1136/bmjresp-2019-000478>.(67)  Royal College of Physicians. National Asthma and Chronic Obstructive  Pulmonary Disease Audit Programme (NACAP) COPD clinical audit 2019/ 2021 June. 2021. [https://www.nacap.org.uk/nacap/welcome.nsf/vwFiles/  COPD+Clinical+Audit+2019-20/$File/NACAP_COPD_SC_Data_And_Methodology_Report_2019-20_Jun_2021.pdf?openelement](74)  European Respiratory Society. An International Comparison of COPD care in Europe, results of the first European COPD audit. 2012 [https://www.ersnet.org/wp-content/uploads/2021/03/copd_audit_web_version.pdf] (75) | **Investment in health and social care**  **Government investment into research**  **Education** | Hall I, Walker S, Holgate ST. Respiratory research in the UK: investing for the next 10 years. Thorax. 2022 Sep 1;77(9):851-3. <http://dx.doi.org/10.1136/thoraxjnl-2021-218459> (52)  Masters R, Anwar E, Collins B, Cookson R, Capewell S. Return on investment of public health interventions: a systematic review. J Epidemiology Community Health. 2017 Aug 1;71(8):827-34. <http://dx.doi.org/10.1136/jech-2016-208141>(59) |
| **Poor provision of care**   - **Poor regional accessibility (Specialist services in London)** | Asthma + Lung UK Failing on the fundamentals report https://cdn.shopify.com/s/files/1/0221/4446/files/COPD_survey.pdf(56) | **Investment in health and social care**  **Government investment into research**  **Local authority investment**  **Public health investment** | Hall I, Walker S, Holgate ST. Respiratory research in the UK: investing for the next 10 years. Thorax. 2022 Sep 1;77(9):851-3. <http://dx.doi.org/10.1136/thoraxjnl-2021-218459> (52)  Masters R, Anwar E, Collins B, Cookson R, Capewell S. Return on investment of public health interventions: a systematic review. J Epidemiology Community Health. 2017 Aug 1;71(8):827-34. <http://dx.doi.org/10.1136/jech-2016-208141>(59) |
| **Norms and attitudes**   - **Limited identification of important phenotypes (e.g. Alpha 1, Bronchiectasis, sleep disorder** | Garudadri S, Woodruff PG. Targeting chronic obstructive pulmonary disease phenotypes, endotypes, and biomarkers. Annals of the American Thoracic Society. 2018 Dec;15(Supplement 4):S234-8. [10.1513/AnnalsATS.201808-533MG](https://doi.org/10.1513/annalsats.201808-533mg)  (76)  McNicholas WT. COPD-OSA overlap syndrome: evolving evidence regarding epidemiology, clinical consequences, and management. Chest. 2017 Dec 1;152(6):1318-26. <https://doi.org/10.1016/j.chest.2017.04.160> (77)  Hurst JR, Elborn JS, De Soyza A. COPD–bronchiectasis overlap syndrome. European Respiratory Journal. 2015 Feb 1;45(2):310-3 10.1183/09031936.00170014 (78) | **Better investment in research** | Hall I, Walker S, Holgate ST. Respiratory research in the UK: investing for the next 10 years. Thorax. 2022 Sep 1;77(9):851-3 <http://dx.doi.org/10.1136/thoraxjnl-2021-218459>. (52)  Masters R, Anwar E, Collins B, Cookson R, Capewell S. Return on investment of public health interventions: a systematic review. J Epidemiology Community Health. 2017 Aug 1;71(8):827-34. <http://dx.doi.org/10.1136/jech-2016-208141>(59) |
| **Poor prevision of care**  **-insufficient clinical staff** | British Thoracic Society. A respiratory workforce for the future. 2022. [https://www.brit-thoracic.org.uk/workforce/] (79) | **Investment in health and social care** | Hall I, Walker S, Holgate ST. Respiratory research in the UK: investing for the next 10 years. Thorax. 2022 Sep 1;77(9):851-3. <http://dx.doi.org/10.1136/thoraxjnl-2021-218459> (52)  Masters R, Anwar E, Collins B, Cookson R, Capewell S. Return on investment of public health interventions: a systematic review. J Epidemiology Community Health. 2017 Aug 1;71(8):827-34 <http://dx.doi.org/10.1136/jech-2016-208141>.(59) |
| **Norms and attitudes**  **-Multimorbidity missed** | Pleasants RA, Riley IL, Mannino DM. Defining and targeting health disparities in chronic obstructive pulmonary disease. International journal of chronic obstructive pulmonary disease. 2016 Oct 4:2475-96 [10.2147/COPD.S79077](https://doi.org/10.2147/COPD.S79077).(80) | **Investment in health and social care**  **Government investment into research**  **Local authority investment**  **Public health investment** | Hall I, Walker S, Holgate ST. Respiratory research in the UK: investing for the next 10 years. Thorax. 2022 Sep 1;77(9):851-3. <http://dx.doi.org/10.1136/thoraxjnl-2021-218459> (52)  Masters R, Anwar E, Collins B, Cookson R, Capewell S. Return on investment of public health interventions: a systematic review. J Epidemiology Community Health. 2017 Aug 1;71(8):827-34 <http://dx.doi.org/10.1136/jech-2016-208141>.(59) |
| **Norms and attitudes**   - **COPD guidelines not implemented** | Sehl J, O’Doherty J, O’Connor R, O’Sullivan B, O’Regan A. Adherence to COPD management guidelines in general practice? A review of the literature. Irish Journal of Medical Science (1971-). 2018 May;187:403-7 [10.1007/s11845-017-1651-7](https://doi.org/10.1007/s11845-017-1651-7). (81) | **Investment in health and social care**  **Fund NHS** | Hall I, Walker S, Holgate ST. Respiratory research in the UK: investing for the next 10 years. Thorax. 2022 Sep 1;77(9):851-3. <http://dx.doi.org/10.1136/thoraxjnl-2021-218459> (52)  Masters R, Anwar E, Collins B, Cookson R, Capewell S. Return on investment of public health interventions: a systematic review. J Epidemiology Community Health. 2017 Aug 1;71(8):827-34. <http://dx.doi.org/10.1136/jech-2016-208141>(59)  Merino M, Martín Lorenzo T, Maravilla-Herrera P, Ancochea J, Gómez Sáenz JT, Hass N, Molina J, Peces-Barba G, Trapero-Bertran M, Trigueros Carrero JA, Hidalgo-Vega Á. A social return on investment analysis of improving the management of chronic obstructive pulmonary disease within the spanish national healthcare system. International Journal of Chronic Obstructive Pulmonary Disease. 2022 Jun 21:1431-42.[[10.2147/COPD.S361700](https://doi.org/10.2147/COPD.S361700)](82) |

**(4) Low status of COPD**

Ways in which both COPD as a condition and people with COPD are held in less regard and considered less of as priority than for other health problems.

Mechanisms and mitigation

| **Mechanisms** | References | **Mitigation** | References |
| --- | --- | --- | --- |
| **Institutional factors**   - **Inadequate research funding- limited advancement in therapies** - **Third sector investment** | Williams S, Sheikh A, Campbell H, Fitch N, Griffiths C, Heyderman RS, Jordan RE, Katikireddi SV, Tsiligianni I, Obasi A. Respiratory research funding is inadequate, inequitable, and a missed opportunity. The Lancet Respiratory Medicine. 2020 Aug 1;8(8):e67-8. <https://doi.org/10.1016/S2213-2600(20)30329-5> (83)  Barnes PJ, Bonini S, Seeger W, Belvisi MG, Ward B, Holmes A. Barriers to new drug development in respiratory disease. European Respiratory Journal. 2015 May 1;45(5):1197-207. 10.1183/09031936.00007915 (84) | **Governmental investment into healthcare**  **Charity sector investment** | Hall I, Walker S, Holgate ST. Respiratory research in the UK: investing for the next 10 years. Thorax. 2022 Sep 1;77(9):851-3. <http://dx.doi.org/10.1136/thoraxjnl-2021-218459> (52)  Masters R, Anwar E, Collins B, Cookson R, Capewell S. Return on investment of public health interventions: a systematic review. J Epidemiology Community Health. 2017 Aug 1;71(8):827-34. <http://dx.doi.org/10.1136/jech-2016-208141>(59) |
| **Institutional factors**   - **Low priorities for commissioners of care** | Asthma and Lung UK. Shaping the future of Respiratory Research and Innovation. 2023. https://www.blog.asthmaandlung.org.uk/blog/future-research-respiratory(85) | **Education**  **Investment**  **Legislation** | Hall I, Walker S, Holgate ST. Respiratory research in the UK: investing for the next 10 years. Thorax. 2022 Sep 1;77(9):851-3. <http://dx.doi.org/10.1136/thoraxjnl-2021-218459> (52)  Masters R, Anwar E, Collins B, Cookson R, Capewell S. Return on investment of public health interventions: a systematic review. J Epidemiology Community Health. 2017 Aug 1;71(8):827-34. <http://dx.doi.org/10.1136/jech-2016-208141>(59) |
| **Institutional factors**   - **Focus on AECOPD to reduce healthcare costs over lived experience** | Asthma + Lung UK Failing on the fundamentals report <https://cdn.shopify.com/s/files/1/0221/4446/files/COPD_survey.pdf> (56) | **Education (medical/nursing school)**  **Mass media**  **Education**  **Governmental investment into healthcare**  **Charity sector investment** | Wakefield MA, Loken B, Hornik RC. Use of mass media campaigns to change health behaviour. The lancet. 2010 Oct 9;376(9748):1261-71.[ <https://doi.org/10.1016/S0140-6736(10)60809-4>] (4)  Masters R, Anwar E, Collins B, Cookson R, Capewell S. Return on investment of public health interventions: a systematic review. J Epidemiology Community Health. 2017 Aug 1;71(8):827-34. <http://dx.doi.org/10.1136/jech-2016-208141>(59)  Hall I, Walker S, Holgate ST. Respiratory research in the UK: investing for the next 10 years. Thorax. 2022 Sep 1;77(9):851-3 <http://dx.doi.org/10.1136/thoraxjnl-2021-218459>.(52) |
| **Norms and attitudes**   - **Historical lack of clinical priority, leading to fewer treatment successes** | Asthma and Lung UK. Shaping the future of Respiratory Research and Innovation. 2023  <https://www.blog.asthmaandlung.org.uk/blog/future-research-respiratory>(85)  Williams S, Sheikh A, Campbell H, Fitch N, Griffiths C, Heyderman RS, Jordan RE, Katikireddi SV, Tsiligianni I, Obasi A. Respiratory research funding is inadequate, inequitable, and a missed opportunity. The Lancet Respiratory Medicine. 2020 Aug 1;8(8):e67-8 <https://doi.org/10.1016/S2213-2600(20)30329-5> (83) | **Governmental investment into healthcare**  **Charity sector investment** | Hall I, Walker S, Holgate ST. Respiratory research in the UK: investing for the next 10 years. Thorax. 2022 Sep 1;77(9):851-3. <http://dx.doi.org/10.1136/thoraxjnl-2021-218459> (52)  Masters R, Anwar E, Collins B, Cookson R, Capewell S. Return on investment of public health interventions: a systematic review. J Epidemiology Community Health. 2017 Aug 1;71(8):827-34. <http://dx.doi.org/10.1136/jech-2016-208141>(59) |
| **Norms and attitudes**   - **Stigma due to link with smoking** | Mathioudakis AG, Ananth S, Vestbo J. Stigma: an unmet public health priority in COPD. The Lancet Respiratory Medicine. 2021 Sep 1;9(9):955-6 <https://doi.org/10.1016/S2213-2600(21)00316-7>.(53)  Walters JA, Hansen EC, Walters EH, Wood-Baker R. Under-diagnosis of chronic obstructive pulmonary disease: a qualitative study in primary care. Respiratory medicine. 2008 May 1;102(5):738-43. <https://doi.org/10.1016/j.rmed.2007.12.008> (66)  Brighton LJ, Chilcot J, Maddocks M. Social dimensions of chronic respiratory disease: stigma, isolation, and loneliness. Current Opinion in Supportive and Palliative Care. 2022 Dec 1;16(4):195-202. <https://doi.org/10.1097/SPC.0000000000000616>(54) | **Education (medical/nursing school)**  **Mass media**  **Education** | Wakefield MA, Loken B, Hornik RC. Use of mass media campaigns to change health behaviour. The lancet. 2010 Oct 9;376(9748):1261-71. [<https://doi.org/10.1016/S0140-6736(10)60809-4>](4)  Masters R, Anwar E, Collins B, Cookson R, Capewell S. Return on investment of public health interventions: a systematic review. J Epidemiology Community Health. 2017 Aug 1;71(8):827-34. <http://dx.doi.org/10.1136/jech-2016-208141>(59) |
| **Norms and attitudes**   - **Limited advancement in novel therapies causing nihilism** | Garudadri S, Woodruff PG. Targeting chronic obstructive pulmonary disease phenotypes, endotypes, and biomarkers. Annals of the American Thoracic Society. 2018 Dec;15(Supplement 4):S234-8. [10.1513/AnnalsATS.201808-533MG](https://doi.org/10.1513/annalsats.201808-533mg)  (76) | **Governmental investment into healthcare**  **Charity sector investment** | Hall I, Walker S, Holgate ST. Respiratory research in the UK: investing for the next 10 years. Thorax. 2022 Sep 1;77(9):851-3 <http://dx.doi.org/10.1136/thoraxjnl-2021-218459> (52)  Masters R, Anwar E, Collins B, Cookson R, Capewell S. Return on investment of public health interventions: a systematic review. J Epidemiology Community Health. 2017 Aug 1;71(8):827-34. <http://dx.doi.org/10.1136/jech-2016-208141>(59) |
| **Norms and attitudes**   - **Patients lacking in social/ cultural capital** | Burney P. Chronic respiratory disease–the acceptable epidemic? Clinical Medicine. 2017 Feb;17(1):29. [10.7861/clinmedicine.17-1-29](https://doi.org/10.7861%2Fclinmedicine.17-1-29) (13) | **Investment in health and social care**  **Reductions in poverty**  **Town planning** | Hall I, Walker S, Holgate ST. Respiratory research in the UK: investing for the next 10 years. Thorax. 2022 Sep 1;77(9):851-3. <http://dx.doi.org/10.1136/thoraxjnl-2021-218459> (52)  Masters R, Anwar E, Collins B, Cookson R, Capewell S. Return on investment of public health interventions: a systematic review. J Epidemiology Community Health. 2017 Aug 1;71(8):827-34. <http://dx.doi.org/10.1136/jech-2016-208141>(59) |
| **Norms and attitudes**   - **Low general awareness of COPD** | Boehm A, Pizzini A, Sonnweber T, Loeffler-Ragg J, Lamina C, Weiss G, Tancevski I. Assessing global COPD awareness with Google Trends. European Respiratory Journal. 2019 Jun 1;53(6) <https://doi.org/10.1183/13993003.00351-2019>.(86) | **Technology**  **Mass media**  **New face of COPD** | Wakefield MA, Loken B, Hornik RC. Use of mass media campaigns to change health behaviour. The lancet. 2010 Oct 9;376(9748):1261-71.[ <https://doi.org/10.1016/S0140-6736(10)60809-4>] (4) |

**(5) Lack of Support**

Factors that make living with COPD more difficult than it should be. Mechanisms and mitigation

| **Mechanisms** | References | **Mitigation** | References |
| --- | --- | --- | --- |
| **Social isolation**  **-transport**  **-local services**  **-recreation** | Metting E, Van Der Molen T, Kocks J. Loneliness and lack of social support severely influences patients' quality of life. Secondary findings from our focusgroup study in asthma and COPD patients <https://doi.org/10.1183/13993003.congress-2016.PA729> .(87) | **Increase social security.**  **Town planning**  **15-minute cities** | Lyu Y, Forsyth A. Planning, aging, and loneliness: reviewing evidence about built environment effects. Journal of planning literature. 2022 Feb;37(1):28-48.  https://doi.org/10.1177/0885412221103513 (88) |
| **Socioenvironmental factors**   - **Inability to continue work.** | Fletcher MJ, Upton J, Taylor-Fishwick J, Buist SA, Jenkins C, Hutton J, Barnes N, Van Der Molen T, Walsh JW, Jones P, Walker S. COPD uncovered: an international survey on the impact of chronic obstructive pulmonary disease [COPD] on a working age population. BMC public health. 2011 Dec;11(1):1-3 [10.1186/1471-2458-11-612](https://doi.org/10.1186/1471-2458-11-612)  .(89) | **Protection for workers**  **Legislation** | Robson LS, Clarke JA, Cullen K, Bielecky A, Severin C, Bigelow PL, Irvin E, Culyer A, Mahood Q. The effectiveness of occupational health and safety management system interventions: a systematic review. Safety science. 2007 Mar 1;45(3):329-53. <https://doi.org/10.1016/j.ssci.2006.07.003> (36) |
| **Socioenvironmental factors**   - **Poor access to advocacy** | Asthma + Lung UK Failing on the fundamentals report https://cdn.shopify.com/s/files/1/0221/4446/files/COPD_survey.pdf(56) | **Invest in healthcare services.**  **Investment in local authority PH funding** | Masters R, Anwar E, Collins B, Cookson R, Capewell S. Return on investment of public health interventions: a systematic review. J Epidemiology Community Health. 2017 Aug 1;71(8):827-34. <http://dx.doi.org/10.1136/jech-2016-208141>(59) |
| **Socioenvironmental factors**   - **Lack of social security e.g. housing, benefits** | Raju S, Keet CA, Paulin LM, Matsui EC, Peng RD, Hansel NN, McCormack MC. Rural residence and poverty are independent risk factors for chronic obstructive pulmonary disease in the United States. American journal of respiratory and critical care medicine. 2019 Apr 15;199(8):961-9 <https://doi.org/10.1164/rccm.201807-1374OC>  (60) | **Reductions in poverty**  **Increase social security.**  **UBI** | Haagh L, Rohregger B. Universal basic income policies and their potential for addressing health inequities: Transformative approaches to a healthy, prosperous life for all. World Health Organization. Regional Office for Europe; 2019. WHO/EURO:2019-3533-43292-60676 (44)  Gibson M, Hearty W, Craig P. The public health effects of interventions similar to basic income: a scoping review. The Lancet Public Health. 2020 Mar 1;5(3):e165-76. [<https://doi.org/10.1016/S2468-2667(20)30005-0>] (5) |
| **Socioenvironmental factors**   - **Digital exclusion from online services.** | Watson A, Wilkinson TM. Digital healthcare in COPD management: a narrative review on the advantages, pitfalls, and need for further research. Therapeutic Advances in Respiratory Disease. 2022 Mar; <https://doi.org/10.1177/17534666221075493>.(90) | **Investment in community services**  **Investment in healthcare services** | Masters R, Anwar E, Collins B, Cookson R, Capewell S. Return on investment of public health interventions: a systematic review. J Epidemiology Community Health. 2017 Aug 1;71(8):827-34. <http://dx.doi.org/10.1136/jech-2016-208141>(59)  Hall I, Walker S, Holgate ST. Respiratory research in the UK: investing for the next 10 years. Thorax. 2022 Sep 1;77(9):851-3. <http://dx.doi.org/10.1136/thoraxjnl-2021-218459> (52) |
| **Socioenvironmental factors**   - **Food, warmth, shelter** | Williams PJ, Cumella A, Philip KE, Laverty AA, Hopkinson NS. Smoking and socioeconomic factors linked to acute exacerbations of COPD: analysis from an Asthma+ Lung UK survey. BMJ Open Respiratory Research. 2022 Jul 1;9(1):e001290 <http://dx.doi.org/10.1136/bmjresp-2022-001290>.(91) | **Reductions in poverty**  **Increase social security**  **UBI** | Haagh L, Rohregger B. Universal basic income policies and their potential for addressing health inequities: Transformative approaches to a healthy, prosperous life for all. World Health Organization. Regional Office for Europe; 2019. WHO/EURO:2019-3533-43292-60676 (44)  Gibson M, Hearty W, Craig P. The public health effects of interventions similar to basic income: a scoping review. The Lancet Public Health. 2020 Mar 1;5(3):e165-76. [I:<https://doi.org/10.1016/S2468-2667(20)30005-0>] (5) |
| **Socioenvironmental factors**   - **Poor access to psychological therapies** | Asthma + Lung UK Failing on the fundamentals report https://cdn.shopify.com/s/files/1/0221/4446/files/COPD_survey.pdf (56)  Pumar MI, Gray CR, Walsh JR, Yang IA, Rolls TA, Ward DL. Anxiety and depression—Important psychological comorbidities of COPD. Journal of thoracic disease. 2014 Nov;6(11):1615. [10.3978/j.issn.2072-1439.2014.09.28](https://doi.org/10.3978%2Fj.issn.2072-1439.2014.09.28) (92) | **Investment in community services**  **Investment in healthcare services**  **Specialist MH services** | Masters R, Anwar E, Collins B, Cookson R, Capewell S. Return on investment of public health interventions: a systematic review. J Epidemiology Community Health. 2017 Aug 1;71(8):827-34. <http://dx.doi.org/10.1136/jech-2016-208141>(59) |
| **Social isolation**   - **Lack of public understanding/ attention/ empathy** | Boehm A, Pizzini A, Sonnweber T, Loeffler-Ragg J, Lamina C, Weiss G, Tancevski I. Assessing global COPD awareness with Google Trends. European Respiratory Journal. 2019 Jun 1;53(6). <https://doi.org/10.1183/13993003.00351-2019> (86) | **Mass media**  **Education**  **Public health investment**  **Few face of COPD** | Wakefield MA, Loken B, Hornik RC. Use of mass media campaigns to change health behaviour. The lancet. 2010 Oct 9;376(9748):1261-71. [<https://doi.org/10.1016/S0140-6736(10)60809-4>] (4)  Hall I, Walker S, Holgate ST. Respiratory research in the UK: investing for the next 10 years. Thorax. 2022 Sep 1;77(9):851-3 <http://dx.doi.org/10.1136/thoraxjnl-2021-218459>.(52) |
| **Social isolation**   - **Lack of peer support networks** | Asthma + Lung UK Failing on the fundamentals report https://cdn.shopify.com/s/files/1/0221/4446/files/COPD_survey.pdf(56) | **Investment in community services**  **Investment in healthcare services** | Masters R, Anwar E, Collins B, Cookson R, Capewell S. Return on investment of public health interventions: a systematic review. J Epidemiology Community Health. 2017 Aug 1;71(8):827-34. <http://dx.doi.org/10.1136/jech-2016-208141> (59)  Hall I, Walker S, Holgate ST. Respiratory research in the UK: investing for the next 10 years. Thorax. 2022 Sep 1;77(9):851-3. <http://dx.doi.org/10.1136/thoraxjnl-2021-218459> (52) |

74.	Royal College of Physicians. National Asthma and Chronic Obstructive Pulmonary Dieasese Audit Programme (NACAP) COPD clinical audit 2019/20. 2021.

75.	European Respiratory Society. An International comprasion of COPD care in Europe, Results of the first European COPD audit. . European Respiratory Society 2012, .

76.	Garudadri S, Woodruff PG. Targeting chronic obstructive pulmonary disease phenotypes, endotypes, and biomarkers. Annals of the American Thoracic Society. 2018;15(Supplement 4):S234-S8.

77.	McNicholas WT. COPD-OSA overlap syndrome: evolving evidence regarding epidemiology, clinical consequences, and management. Chest. 2017;152(6):1318-26.

78.	Hurst JR, Elborn JS, De Soyza A. COPD–bronchiectasis overlap syndrome. Eur Respiratory Soc; 2015. p. 310-3.

79.	British Thoracic Society. A respiratory workforce for the future. 2022.

87.	Metting E, Van Der Molen T, Kocks J. Loneliness and lack of social support severely influences patients' quality of life. Secondary findings from our focusgroup study in asthma and COPD patients. Eur Respiratory Soc; 2016.
